# Therapeutic Targeting of Oncogene-induced Transcription-Replication Conflicts in Pancreatic Ductal Adenocarcinoma

**DOI:** 10.1101/2023.10.03.23296487

**Authors:** Shanna J. Smith, Fan Meng, Robert G. Lingeman, Caroline M. Li, Min Li, Galyah Boneh, Toni T. Seppälä, Thuy Phan, Haiqing Li, Richard A Burkhart, Vishwas Parekh, Syed Rahmanuddin, Laleh G. Melstrom, Robert J. Hickey, Vincent Chung, Yilun Liu, Linda H. Malkas, Mustafa Raoof

## Abstract

**Purpose:** Transcription-replication conflicts (TRCs) are a major source of endogenous replication stress in cancer. We previously discovered that pancreatic ductal adenocarcinoma (PDAC) demonstrates uniquely high levels of TRCs compared to other common solid tumors. Here, we characterize the mechanism of action, oncogene-dependency, PDAC subtype-specificity, and preclinical activity of a TRC-targeting small molecule – AOH1996 – in a spectrum of PDAC models. We also provide first clinical evidence of the activity of AOH1996 in a PDAC patient.

**Experimental Design:** The oncogene-dependent toxicity of AOH1996 was examined in KRAS(G12D) inducible systems. Next, the effect of AOH1996 was evaluated on replication fork progression, TRCs, DNA damage response, cell cycle progression, and apoptosis in PDAC cell lines. PDAC subtype-specific responses were evaluated in organoid cultures, and *in vivo* efficacy was evaluated in murine and patient-derived xenografts. Efficacy in a PDAC patient was evaluated by radiographic response assessment and progression-free survival.

**Results:** AOH1996 demonstrated dose-dependent cytotoxicity that was exquisitely dependent on oncogenic KRAS(G12D) induction. Cytotoxicity of AOH1996 was evident in several human and murine PDAC cell lines (Average IC50 across cell lines 0.72μM). Mechanistically, AOH1996 inhibited replication fork progression and promoted TRCs through enhanced interaction between RNA Polymerase II and Proliferating cell nuclear antigen which resulted in transcription-dependent DNA damage and global transcription shutdown. AOH1996 demonstrated activity in all organoid lines tested with varying potency (IC50 406nM – 2μM). Gene expression analysis demonstrated that organoids with replication stress high or very strongly basal signature were most vulnerable to AOH1996. In PDAC mouse model studies, AOH1996 reduced tumor growth rate, enhanced tumor-selective DNA damage and prolonged survival (Median 14 days vs. 21 days, P=0.04) without observable toxicity. The first patient with chemotherapy-refractory PDAC who was treated with AOH1996 monotherapy demonstrated early evidence of efficacy (49% shrinkage of the two hepatic metastases with stabilization of disease at other sites).

**Conclusions:** Therapeutic targeting of TRCs using small molecule inhibition is safe and effective in preclinical models. Pre-clinical data along with proof-of-concept activity in a patient with chemotherapy-refractory PDAC provides rationale for further clinical development of TRC targeting strategies.

## INTRODUCTION

Genomic instability in cancers is a direct consequence of DNA damage from both endogenous and exogenous insults (1). Replication stress from oncogenes - such as KRAS and MYC in PDAC – is a major source of endogenous insult on the DNA. On a molecular scale, oncogenes place significant demands on the DNA replication machinery such that errors in the process cannot be rectified (2). The oncogene – replication stress – genomic instability axis is now a well-accepted hallmark of oncogene-driven cancers such as PDAC (3).

Oncogenic KRAS is a pathognomonic feature of PDAC, occurring in 95% of patients; with KRAS(G12D) alteration comprising half the mutations (4). Oncogenic KRAS in PDAC can drive DNA damage through production of reactive oxygen species, as well as hyper-replication (5). Recent studies have implicated hyper-transcription as another major impediment to replication fork progression causing DNA damage (2,6,7). To cope with the excessive DNA damage, KRAS-driven PDAC cells activate DNA damage response pathways such as ATR-Chk1 to allow DNA repair. At the same time inactivation of apoptotic pathways, through co-mutation in TP53, permits DNA damage tolerance in the face of excessive replication stress.

Therapeutic strategies against KRAS-driven genome instability may either involve inhibiting oncogenic signaling to minimize DNA damage; or inhibiting replication stress-triggered adaptive mechanisms, thereby promoting lethal DNA damage. Until recently, direct targeting of most oncogenic KRAS mutations was not possible (8). Early attempts at therapeutic targeting of DNA repair pathways (e.g., Chk1 inhibition) were also met with disappointing results in the clinic (9). Recently, however, FDA approved a first-in-class poly(adenosine diphosphate–ribose) polymerase (PARP) inhibitor which produces synthetically lethal DNA damage in tumors with homologous recombination (HR) DNA repair pathway deficiency (10). BRCA genes involved in the HR pathway have a germline mutation in approximately 4-7% of individuals with PDAC (11). In these patients, compared to placebo, PARP inhibitor treatment resulted in a significantly longer progression-free survival (7.4 months vs. 3.8 months; hazard ratio for disease progression or death, 0.53; 95% confidence interval [CI], 0.35 to 0.82; P=0.004) (12,13). This landmark phase 3 clinical trial supports the notion that targeted, synthetically lethal DNA-damaging therapies can improve the dismal outcomes of individuals with PDAC (14).

Most individuals with PDAC do not have HR deficiency. Therefore, novel strategies that promote cancer-specific DNA damage are needed. Based on transcriptomic analysis, PDAC is now classified into two main subtypes: classical and basal (15). The basal subtype is associated with therapy resistance; and worse survival (16,17). A deeper analysis of the transcriptomic subtypes demonstrates that the genes associated with replication stress are enriched in the basal subtype (18). This high replication stress phenotype is distinct from a previously recognized DNA repair deficiency phenotype and may represent a novel therapeutic vulnerability.

In this study, we perform pre-clinical experiments to test the utility of targeting replication stress in PDAC using a recently described small molecule, a bioavailable inhibitor of Proliferating Cell Nuclear Antigen (PCNA) – AOH1996 (19). PCNA is known as the ringmaster of the genome (20). Evolutionarily, it is highly conserved because of its central role in orchestrating DNA replication and repair. More specifically, it forms a homotrimer processivity clamp encircling the DNA strand. This orientation with respect to the DNA allows PCNA to act as a scaffold for binding of key proteins during DNA replication and repair. Although, these functions of PCNA are ubiquitous to all proliferating cells, their requirement in cancer cell survival is particularly critical. Previous studies on human tissues demonstrate that PCNA is highly expressed in PDAC compared to adjacent normal tissues (21). Further, PCNA expression correlates with tumor cell differentiation and lymph node metastases, suggesting that PCNA plays a key role in pathogenesis of aggressive PDAC (22).

Efforts at targeting PCNA were previously limited because of concerns of toxicity of PCNA inhibitors toward normal tissues (23). However, recent development of non-toxic cancer-selective PCNA inhibitor – AOH1996 – provides an opportunity to target basal (replication stress high) PDAC (19). Here, we characterize the mechanism of action, oncogene-dependency, PDAC subtype-specificity, and activity of AOH1996 in a spectrum of preclinical PDAC models, including; cell lines, organoids, and murine cancer models. Further, we present the first evidence of clinical activity of AOH1996 in a patient with PDAC. These findings lay the foundation for further clinical trials focused on therapeutic targeting of replication stress in PDAC using AOH1996.

## RESULTS

### AOH1996 causes oncogene-dependent toxicity

We first examined the effect of AOH1996 on replication stress phenotype in PDAC. We used a commercially sourced telomerase-immortalized human pancreatic ductal-derived (HPNE) cell line (as previously published (24)). This cell line expresses human papillomavirus E6 and E7 oncogenes, which block the function of the p53 and Rb tumor suppressors, respectively. In addition, SV40 small t antigen is expressed as it is required to allow oncogenic transformation upon KRAS(G12D) expression. We have previously demonstrated that, upon doxycycline induction, this system causes replication stress, R-loops, transcription-replication conflicts and DNA damage (7). We exposed the HPNE cells with doxycycline-inducible oncogenic KRAS(G12D) to AOH1996 or DMSO, with or without KRAS(G12D) induction. This experiment demonstrated that AOH1996 selectively induced DNA damage – as measured by γH2AX induction – in HPNE cells expressing oncogenic KRAS but not in control conditions (Figure 1A)

**Figure 1.**
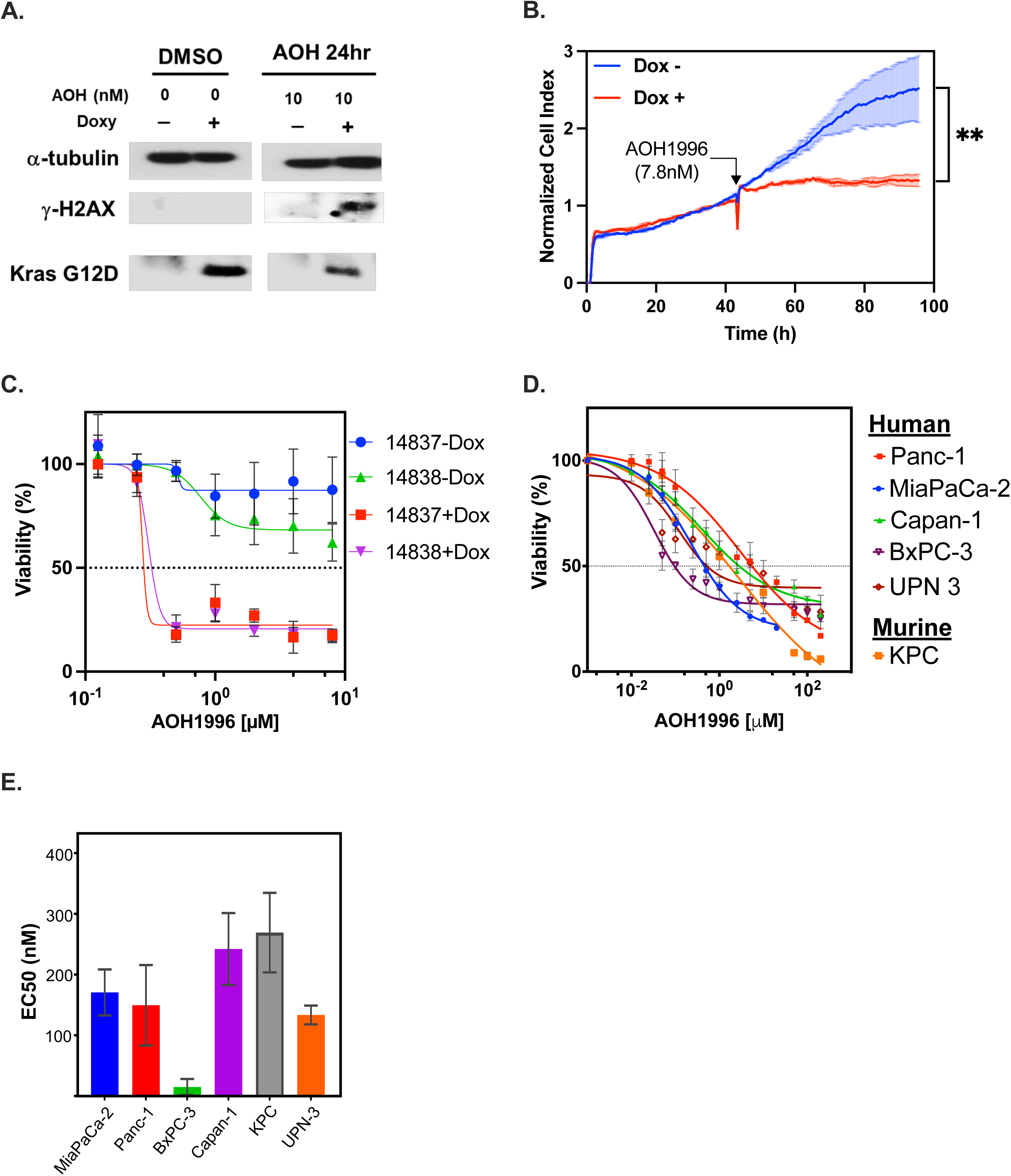
AOH1996 causes oncogene-dependent toxicity. **A.** Representative **western blot** of the indicated proteins is shown from whole cell lysates of HPNE cells indicating induction of oncogenic KRAS and DNA damage (γH2AX) upon KRAS induction. α-tubulin is used as a loading control. **B. Real-Time Cell Analysis** of HPNE cells with stable expression of doxycycline inducible-KRAS(G12D) vector. Cell-index is a unitless measure of cellular impedance of electron flow caused by adherent cells depicting cell growth and confluence. Lines indicate average of three replicates and error bars indicate standard deviation. **Welch’s t-test comparing cell index at 96h time point: p= 0.009 **C & D.** Exponentially growing cells were exposed to increasing concentrations (0-20 µM) of AOH1996 for 48 hours, and then analyzed using the CellTiter Glo^TM^ assay. **Cell viability** is reported as % of Control, with 100% representing a zero response. Data points indicate mean and error bars indicate standard deviation calculated from at least 3 replicates. **E. Half maximal effective concentration (EC50)** was calculated using Prism. The values represent average and corresponding error bars indicate standard errors determined from 4 separate biological replicates.

To further investigate if AOH1996-induced genotoxicity impacts growth kinetics in an oncogenic KRAS-dependent manner, we performed real-time cell assay (RTCA) that measures cell confluence/ attachment over time. HPNE KRAS(G12D) cells were pre-induced for 72 hours with doxycycline or DMSO, then plated on RTCA plates for tracking the cell proliferation by detecting cell attachment on a sensor. The cells were then allowed to grow, and AOH1996 (7.8nM) was added to both conditions after 40 hours. The initial growth during the first 40 hours was similar in both doxycycline and DMSO conditions. After exposure to AOH1996, there was significant growth inhibition in doxycycline-induced cells, whereas the DMSO-treated cells continued to proliferate at a normal growth trajectory (Figure 1B). For orthogonal validation of these findings, we used murine iKRAS cell lines (25). These cell lines (14837 and 14838) are derived from iKRAS mouse where oncogenic KRAS(G12D) is expressed under doxycycline induction. Withdrawal of doxycycline leads to KRAS(G12D) extinction within 24 hours. iKRAS 14837 and 14838 cells were treated with increasing concentrations of AOH1996 with or without KRAS(G12D) extinction (Figure 1C). Oncogenic KRAS(G12D) expressing cell lines demonstrated exquisite sensitivity to AOH1996 whereas KRAS(G12D) extinct cells were resistant, as measured by Celltiter Glo^TM^ assay. To determine if AOH1996 has activity in PDAC cell lines, we examined cell viability using Celltiter Glo^TM^ assay across a panel of human and murine PDAC cells. The assay demonstrated a robust dose dependent toxicity in PDAC cells with IC50 values ranging from 0.03μM for BxPC3 cells to 2.2μM for Panc1 cells (Average IC50 across cell lines 0.72μM) after 48 hours of exposure to AOH1996 (Figure 1D and E). Notably, BxPC3 cells are KRAS wildtype but have a BRAF mutation. The sensitivity of BxPC3 to AOH1996 suggests that the effect is not limited to mutant KRAS cell lines. Replication stress enhances dependence on homologous recombination mediated mechanisms for replication for fork restart and repair. However, BRCA2-mutant (BRCA2.6174delT) Capan-1 cells demonstrated similar sensitivity to AOH1996 compared to BRCA2-wildtype cell lines (Figure 1D); or compared to BRCA2-revertent Capan-1 C2-14 cells that have restored BRCA2 function through a secondary mutation (Supplementary Figure 1). Together, these data demonstrate that AOH1996 causes oncogene-dependent growth inhibition and cytotoxicity independent of homologous recombination repair proficiency.

### AOH1996 enhances replication stress and DNA damage in PDAC cells

To investigate the mechanism of AOH1996-dependent cytotoxicity, we focused on replication fork dynamics given the central role of PCNA in replication fork progression (20). PDAC cells (MIA Paca-2) were exposed to increasing concentrations of AOH1996, and replication fork progression was measured using DNA fiber analysis. To establish baseline replication fork progression, DNA was labeled (with CldU) for 15 minutes prior to the introduction of AOH1996 or DMSO (control). CldU was washed, and a second label (IdU) was added to track replication fork progression in the presence of AOH1996 (or DMSO). As shown in Figure 2A-B and S2A-B, AOH1996 exhibits a dose-dependent decrease in replication fork progression after normalizing for pre-treatment replication rate. The observed replication fork stalling did not increase the licensing of new origins as a compensatory mechanism (Figure 2C), as there was no statistically significant increase in the number of new origins.

**Figure 2.**
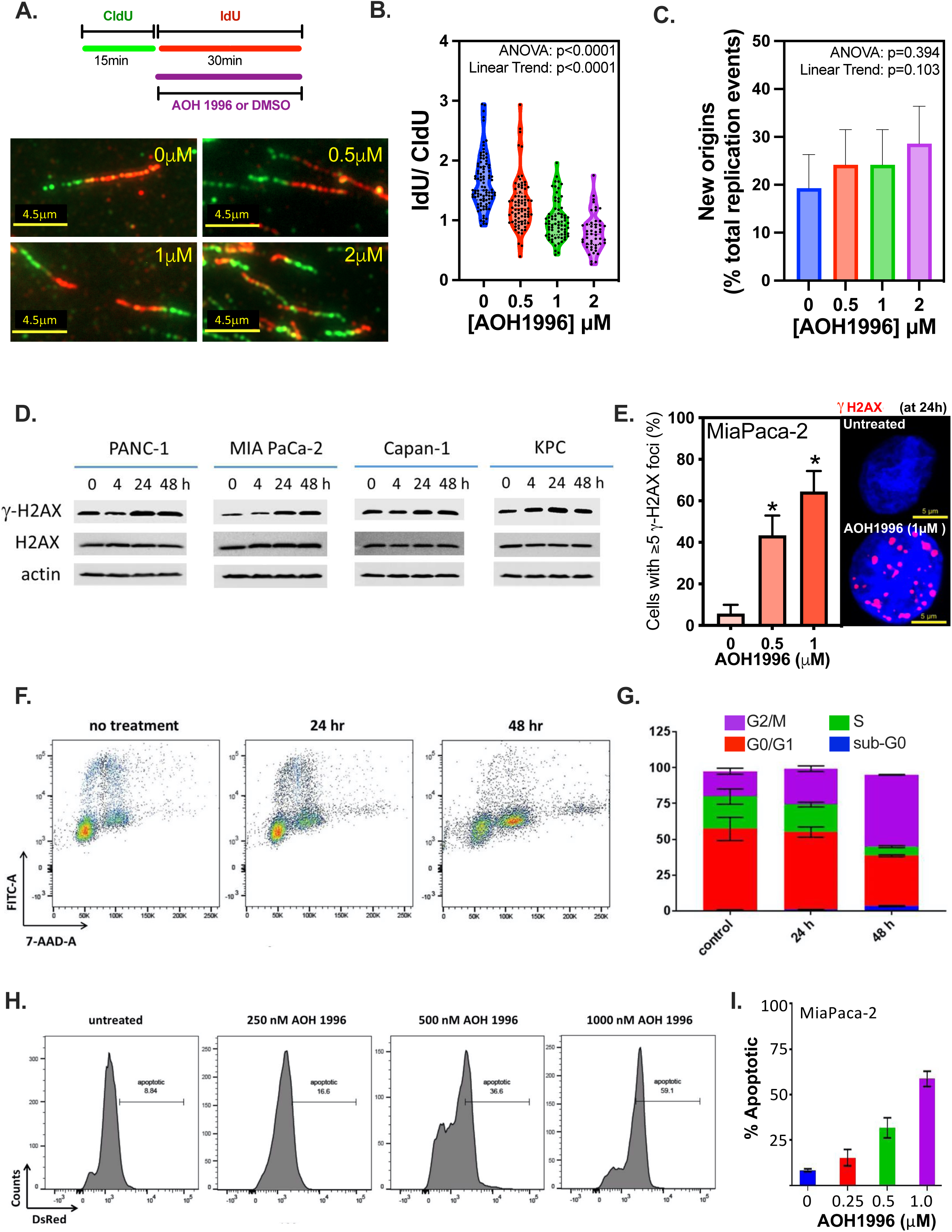
AOH1996 enhances replication stress and DNA damage in PDAC cells. **A. DNA fiber analysis** was performed. Replicating DNA was labeled with CldU for 15min, followed by IdU for 30min. AOH1996 or DMSO was added with IdU labeling. Representative images from four different conditions are shown. **B. Replication fork progression:** IdU tract lengths were normalized to average CldU length prior to drug treatment. Violin plots are shown. One-way ANOVA demonstrated significant differences between the groups (p<0.0001). There was linear trend with increasing dose of AOH1996 (Slope -0.29, p<0.0001). (See Supplemental Figure S2A and S2B for replication speed before and after drug treatment). **C. New Origins:** IdU tracts without preceding CldU tracts were counted and expressed as a proportion of total replication forks (CldU and IdU) to calculate the number of new origins. Error-bars represent 95% confidence interval encompassing proportion estimates. One-way ANOVA did not demonstrate any significant differences between treatments and there was a lack of linear trend, as shown. **D. Western Blot:** Cells were treated with 200 nM AOH1996 for increasing amounts of time (0-48 hours). Representative immunoblots of protein extracts from each pancreatic cancer cell line tested are shown. Actin was used as the loading control. **E. Immunocytochemistry** for DNA damage marker (γH2AX) demonstrated a significant dose-dependent increase in cells treated with AOH1996 compared to controls (A representative experiment is shown. One-way ANOVA with Dunnett adjustment for multiple comparisons, *p<0.0001 relative to control. Error bars represent 95% confident intervals). (Also see Supplemental, Figure S2C). **F & G. Flow cytometry** (BrdU-FITC and 7-AAD) cell cycle analysis. MIA PaCa-2 cells were treated with 200 nM of AOH1996 for 24 or 48 hours, and then analyzed in triplicates. Average proportions are plotted in stacked bar graph with error bars indicating standard deviation. (Dose response is demonstrated in Supplement, Figure S2D). **H & I.** MIA PaCa-2 cells were treated with increasing concentrations of AOH1996 (0-1000 nM) for 24 hours. Apoptotic cells were labeled using **TUNEL assay** and quantified using flow cytometry in triplicates. Average proportions are plotted in bar graph with error bars indicating standard deviation.

Next, we examined if AOH1996-induced inhibition of replication fork progression results in DNA damage in PDAC cells. The panel of pancreatic cancer cell lines were treated with 200 nM AOH1996 up to 48 hours. Cells were harvested at specific time points (0, 4, 24, 48 hours) to be analyzed by Western blot analysis. As Figure 2D illustrates, AOH1996 induces a DNA damage response (γ-H2AX phosphorylation) in all the cell lines tested. To validate these results, MIAPaCa-2 cells were treated with increasing concentrations of AOH1996 (0 – 1000 nM) for 24 hours, and DNA damage was assessed using immunocytochemistry (Figure 2E and S2C). There was significant dose-dependent increase in γ-H2AX levels in the nuclei of AOH1996-treated cells, consistent with the whole cell lysate western blot results.

To evaluate if AOH1996-induced DNA damage causes cell cycle arrest and/ or apoptosis, we evaluated the cell cycle dynamics using flow cytometry. AOH1996 induced a robust late S and G2/M arrest in exponentially growing MIA Paca-2 cells after exposure to low doses (200nM) of AOH1996 for 48 hours (Figure 2F-G), an effect that was more pronounced at higher doses (Supplementary Figure S2D). To determine if this AOH1996-induced genotoxicity and cell cycle arrest leads to apoptotic cell death, MIA PaCa-2 cells were treated with increasing concentrations (0 – 1000 nM) of AOH1996 for 24 hours, then analyzed using TUNEL assay. As illustrated in Figure 2H-I, fragmentation increases with increasing concentrations of AOH1996, thus indicating an increase of apoptotic cells in a dose-dependent manner. Collectively, these findings demonstrate that AOH1996 causes dose-dependent inhibition of replication fork progression, DNA damage and cell death in PDAC cells, in part through apoptosis.

### Impact of AOH1996 on Transcription-replication conflicts (TRCs) in PDAC

We have previously demonstrated that transcription-dependent replication stress through transcription-replication conflicts (TRCs) is a major mechanism of endogenous replication stress in human PDACs (7). TRCs may therefore represent a unique targetable vulnerability in human PDAC. One of the proposed mechanisms of action of AOH1996 is the targeting of TRCs (19). Specifically, AOH1996 promotes an interaction between RNA Polymerase II (APIM motif) and PCNA, potentially increasing TRCs. We examined the impact of AOH1996 on TRCs in Panc1 cells through a flow cytometry-based proximity ligation assay and specifically measured foci with <40nm proximity of RNAPII and PCNA. At baseline, 50% of PDAC cells demonstrated high TRCs. Upon exposure to AOH1996 for 24 hours, this proportion increased to 65-82% depending on the dose of AOH1996 (Figure 3A and B).

**Figure 3.**
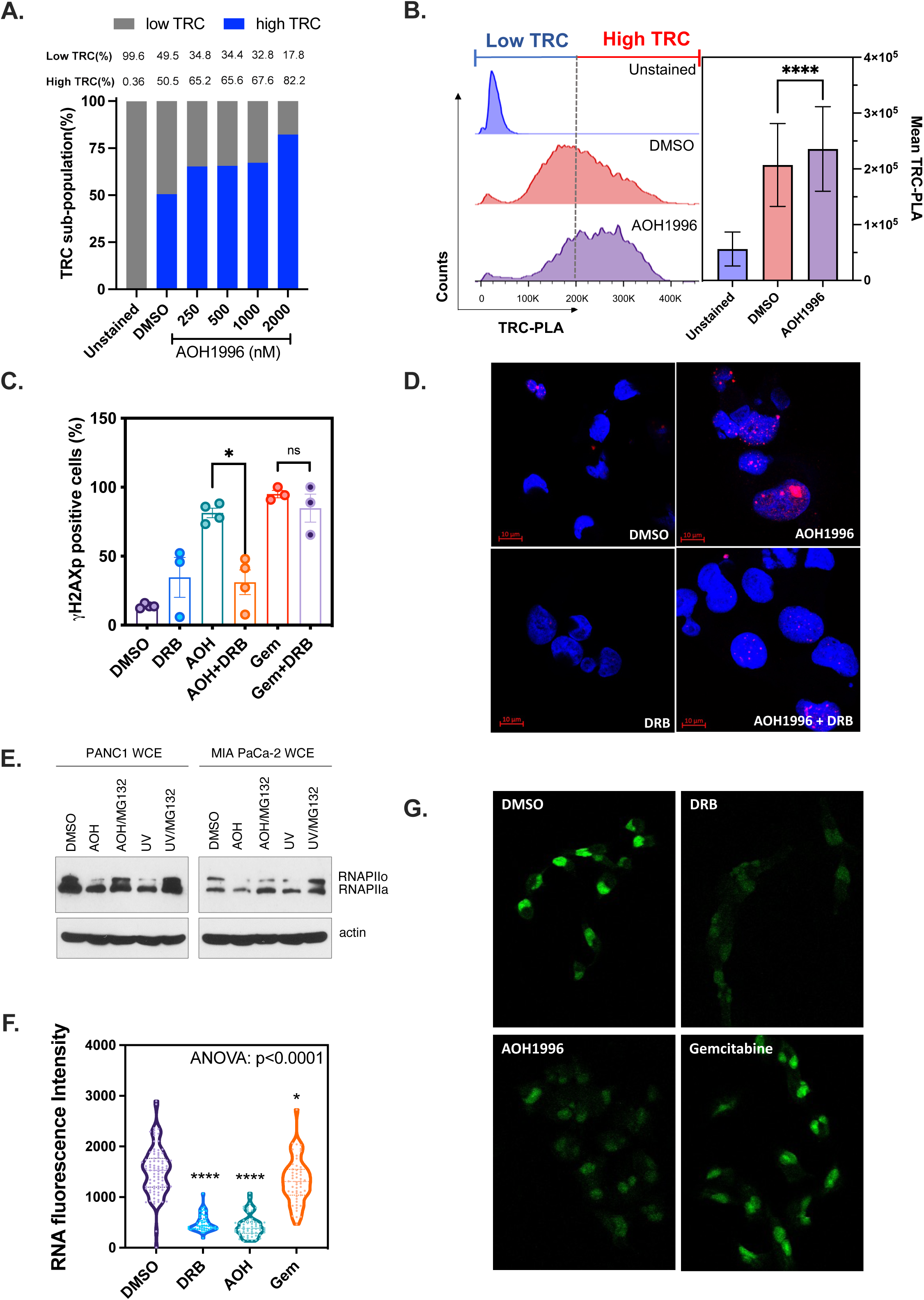
Impact of AOH1996 on Transcription-replication conflicts (TRCs) in PDAC. **A & B. Flow cytometric RNAPII-PCNA Proximity Ligation Assay** was performed in Panc1 cells to quantify the interaction between transcription machinery protein (RNAPII) and replication scaffold (PCNA) as a measure of TRCs. The threshold of high vs. low TRCs was set at the median of control. Representative data are shown in a stacked bar graph A and histogram B. (****p<0.0001, Unpaired t-test). **C & D. Immunocytochemistry** for DNA damage marker (γH2AX) with or without 24h exposure to 500nM AOH1996 or 500nM Gemcitabine or DMSO in MIA PaCa-2 cells. Transcription inhibitor DRB (100μM, last 2 hours) was used as indicated. The data points demonstrate an average of 3-4 independent experiments and error bars indicate standard error. (*p=0.028, Mann-Whitney test.) **E. Western Blot:** Indicated PDAC cells were treated with 500 nM AOH1996 for 12 hours. (0-48 hours). Representative immunoblots of protein extracts from each pancreatic cancer cell line tested are shown. Actin was used as the loading control. UV dose was 30 J/m2 over 30 minutes. **F & G. Global transcription** was measured in MIA Paca-2 cells after exposure to AOH1996 500nM or Gemcitabine 500nM for 12 hours. DRB is used as a positive control for transcription inhibition (100μM, 2 hours). Violin plots depicting fluorescence intensity from a representative experiment are shown in F and images are shown in G. Analysis shown is one-way ANOVA with Dunnett multiple comparison test, comparing different conditions to DMSO control. (****p<0.0001, *0.012)

We then asked if the DNA damage induced by AOH1996 is in part explained by AOH1996-induced increase in TRCs. The exponentially growing MIA Paca-2 cell culture was exposed to AOH1996, with or without a transcription inhibitor – DRB (5,6-dichloro-1-beta-D-ribofuranosylbenzimidazole), – and DNA damage was measured by γ-H2AX immunocytochemistry (Figure 3C and D). The results were compared to DMSO control, gemcitabine treatment, or a combination of gemcitabine and DRB treatment. Gemcitabine, a DNA damaging nucleoside analogue FDA-approved for pancreatic cancer patients, causes transcription-independent DNA damage by stalling of replication forks. The results demonstrate that both AOH1996 and gemcitabine caused DNA damage. But, contrary to gemcitabine, AOH1996-induced DNA damage was significantly inhibited by DRB (81% vs. 31%, p = 0.0286), suggesting that AOH1996-induced DNA damage is at least partially transcription-dependent.

Prior work has demonstrated that persistent TRCs ultimately result in RNAPII degradation to resolve replication stress (26,27). To test this possibility, we evaluated the effects of AOH1996 on RNAPII expression using western blot analysis in pancreatic cancer whole cell lysates (using anti-RNAPII A10 antibody). Our results (Figure 3E) indicate that AOH1996 (500nM for 12 hours) caused degradation of the active RNAPII – RNAPIIo (as opposed to inactive RNAPII – RNAPIIa), which could be inhibited by the proteasome inhibitor, MG132; like the UV exposure (positive control). We then asked if degradation of RNAPIIo would impact overall transcription. As shown in Figure 3F and G, MIA Paca-2 cells exposed to AOH1996 (500nM, 12h) demonstrated a significant decrease in global mRNA synthesis compared to DMSO control, as measured by 5-Ethynyl Uridine (EU) incorporation assay. Transcription inhibitor, DRB, was used as a positive control. Gemcitabine had a very small effect on mRNA synthesis. Collectively, the results from these experiments provide evidence of TRC-dependent DNA damage and transcription shutdown in PDAC cells from treatment with AOH1996.

### AOH1996 targets replication stress-high subtype of PDAC

Considering the limitation of 2D cell culture models, we tested efficacy of AOH1996 in established PDAC organoid cell cultures. AOH1996 demonstrated robust activity in all organoid lines tested with varying potency (IC50 406nM – 2μM) as shown in Figure 4A.

**Figure 4.**
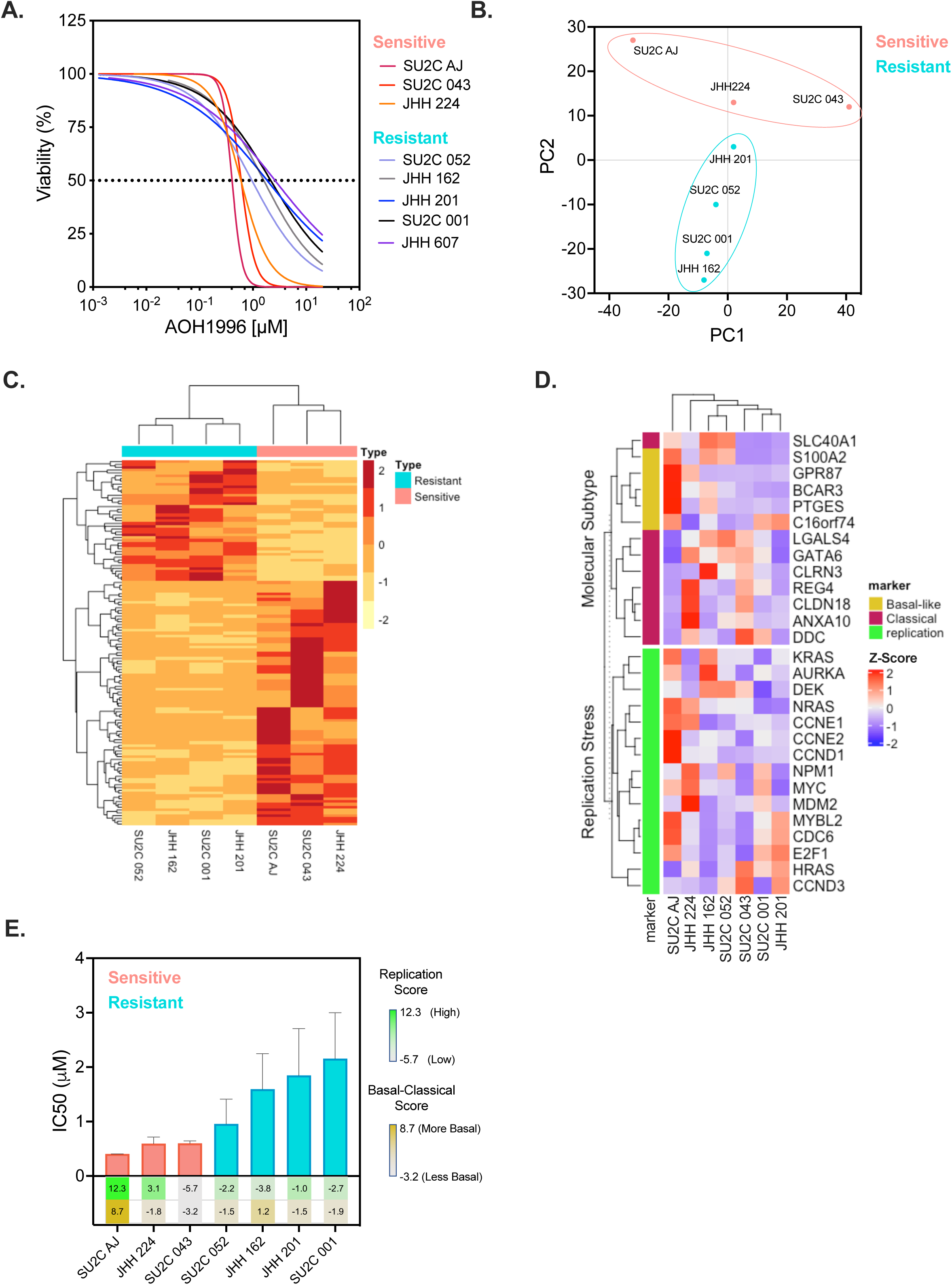
AOH1996 targets replication stress-high subtype of PDAC. **A. PDAC organoids** from untreated patient tumors were exposed to increasing concentrations (0-20 µM) of AOH1996 for 72 hours, and then analyzed using the CellTiter Glo^TM^ assay. **Cell viability** is reported as % of Control, with 100% representing a zero response. Lines indicate best-fit values derived from a log-inhibitor vs. response model with variable slope using Prism. **B. Principal component analysis** derived from gene expression data from the indicated organoid lines. PC1 and PC2 are shown which described most of the variation in the data. **C. Heatmap** of gene expression profiles of sensitive and resistant organoid lines cluster distinctly. 130 top differentially-expressed genes are shown. **D. Gene-expression signatures for PDAC subtypes** are demonstrated for each sample **E.** IC50 values derived from the model in 4A are shown with replication stress and basal signature scores for DNA damage marker (γH2AX) with or without 24h exposure to 500nM.

Recent work has demonstrated that replication stress is a pathognomonic feature of basal subtype of PDAC (18). To determine if AOH1996 toxicity has subtype specificity, we correlated transcriptomic signatures to AOH1996 response. Gene expression analysis was available for 3 drug sensitive and 4 drug resistant organoid lines. A total of 13,279 genes passed the quality filter and were included in the analysis. Principal component analysis (PCA) plot (Figure 4B) and gene expression heatmap (Figure 4C) demonstrate that sensitive and resistant lines cluster separately based on gene expression profiles. There are 130 differentially expressed genes – 43 downregulated genes (drug sensitive vs drug resistant) and 87 upregulated genes (figure 4C). Supplemental Figure 3 highlights the genes that are significantly differentially expressed (Adjusted p-value <0.05). The top 20 differentially expressed genes based on the adjusted p-value are labeled.

Distinct clustering of resistant and sensitive organoid lines based on gene expression analysis suggests that transcriptomic profiles of PDAC predicts sensitivity to AOH1996. The PDAC cell lines were therefore annotated as classical, basal, both, or none based on their gene-expression profiles as previously described (28). SU2C-AJ was strongly basal, whereas JHH-201 was weakly basal. JHH-224 and SU2C-043 demonstrated moderately classical profiles, whereas SU2C-052 was weakly classical. JHH162 demonstrated both basal and classical signatures whereas SU2C-01 demonstrated neither (Figure 4D). Two cell lines were classified as replication stress high, SU2C-AJ and JHH224. AOH1996 was most potent in two out of three cell lines with replication stress high transcriptomic signature (Figure 4E). One of these (SU2C-AJ) also had a strong basal signature whereas the other two did not. Taken together, these data suggest that tumors with replication stress high transcriptomic signature or very strongly basal signature may be vulnerable to AOH1996.

### *In vivo* therapeutic efficacy of AOH1996 in PDAC

To test the efficacy of AOH1996 *in vivo*, we used a fast-growing orthotopic model of pancreatic cancer (average time from implantation to death ∼ 4 weeks). A murine pancreatic cancer cell line isolated from a KPC mouse was used and injected in the body of the pancreas, as described in the “Materials and Methods” section (Figure 6a). After tumor formation, mice were randomized to treatment with AOH1996 at 100 mg/kg or Control (excipients) only. The treatments were administered via oral gavage once a day for 4 consecutive days. The mice were euthanized on day 5 (Figure 5A). Mice treated with AOH1996 had significantly smaller tumors compared to control group (Figure 5C, Supplemental Figure S4). Mice in both groups behaved similarly during the experiment. The mouse weights during the study did not differ between the two groups (Figure 5B). A detailed analysis of the H&E specimens by the pathologist did not demonstrate any histologic evidence of toxicity in intestine, lung, heart, liver, or kidney (Supplemental Figure S5). Tumor samples from the mice treated with AOH1996 had significantly higher proportions of nuclei with DNA damage (γH2AX) compared to control mice (Mean 45.5% vs. 11.8%, Nested t-test, p-value <0.0001, Figure 5D). There was a slight decrease in proliferation rate (measured by Ki67) and slight increase in apoptosis (measured by TUNEL assay), as shown in Supplemental Figure S6. These differences, however, were not statistically significant. The Ki67 and TUNEL positive cells in the spleen were similar in the control and AOH1996 treated mice (Supplemental Figure S6). These findings demonstrate the efficacy of AOH1996 in a murine model of pancreatic cancer without evidence of toxicity to normal tissues over a short duration. We validated the results from this experiment and tested the efficacy of AOH1996 in a patient-derived xenograft model of PDAC, developed from a chemotherapy refractory tumor with KRAS(G12V) and TP53 (p.Tyr205_Leu206delinsTer) co-mutation. AOH1996 significantly delayed the growth of the tumor, like in our previous results (p=0.013, Figure 5E).

**Figure 5.**
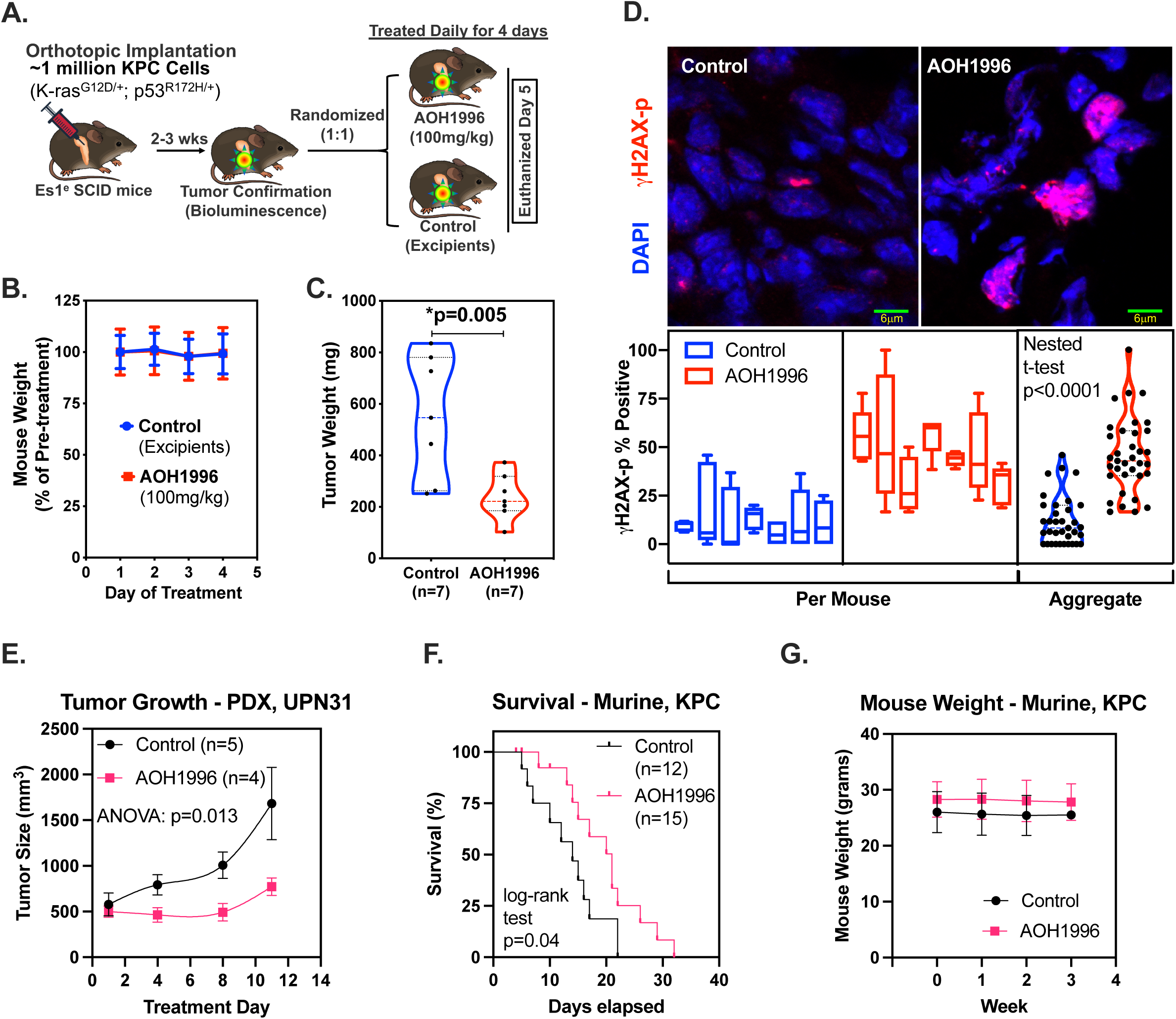
*In vivo* therapeutic efficacy of AOH1996 in PDAC. **A. Mouse Model:** Mouse pancreatic cancer (KPC) cell line was isolated from KPC (LSL-KrasG12D/+; LSL-Trp53R172H/+) mouse PDAC tumor and modified to express luciferase. KPC cells were orthotopically implanted in carboxyl esterase deficient (ES1^e^) SCID mice. Mice were then imaged using bioluminescence at 2-3 weeks after tumor cell implantation to confirm tumor formation and randomized to treatment with AOH1996 or Control. Treatment was given for 4 days. Mice were euthanized on day 5. Mouse (**B**) and Tumor (**C**) weights are shown. (One-sided Mann Whitney test, p=0.005). See Supplemental Figures S4, S5, S6 for additional details. **D. Fluorescent immunostaining** for DNA damage marker (γH2AXp) in fresh frozen sections of tumors treated with AOH1996 compared to controls from experiment 5A (Nested t-test, p<0.0001). At least 5 sections per mouse tumor and 100 cell nuclei were analyzed. **E. Ectopic patient-derived xenograft UPN31** implanted in the flank of carboxyl esterase-deficient ES1^e^ SCID mice were allowed to grow to approximately 500 mm^3^ then randomized to receive AOH1996 (100mg/kg) orally or excipients (Control). Tumor volume was recorded. Mean and standard deviation are reported. **F & G. Survival Experiment.** Mice corresponding to the tumor model in **5A** were randomized after tumor confirmation on bioluminescence and treated with AOH1996 (100 mg/kg, 5 days a week) or excipients (Control) by oral gavage. Mice were euthanized when they met IACUC approved euthanasia criteria. Kaplan-Meier Survival analysis is shown in **F** and tumor weights in **G**.

**Figure 6.**
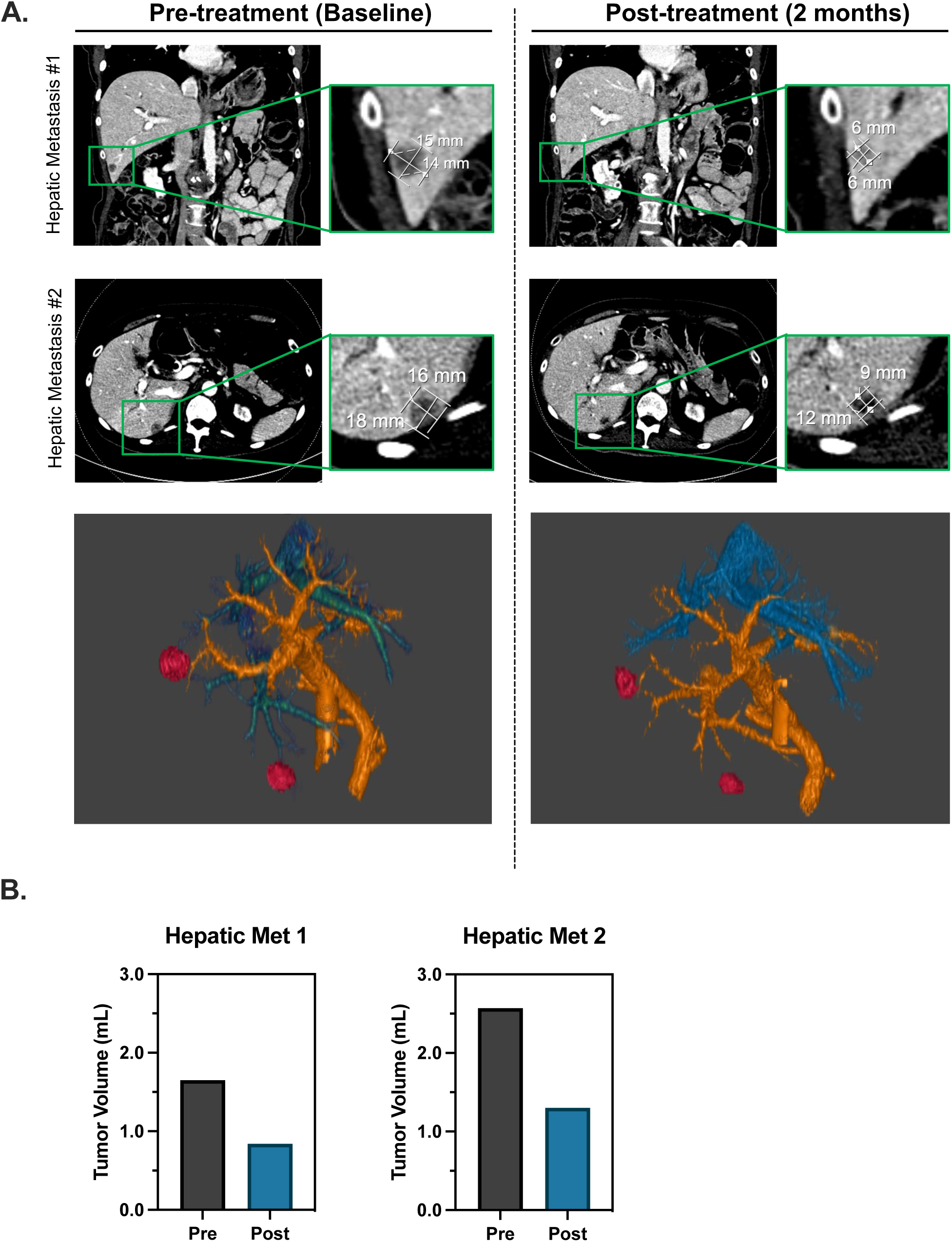
Axial and Coronal images from Computed Tomography scans pre- and post-treatment with AOH1996 from a patient with metastatic PDAC are shown along with 3D reconstruction (**A**) and tumor volume quantification (**B**).

To measure the impact of AOH1996 on survival, we randomized mice with orthotopic KPC tumor to treatment with AOH1996 at 100 mg/kg or Control (excipients only) 2 weeks after orthotopic tumor implantation. Treatments were repeated daily (5 days/week) till the mice were moribund or met euthanasia criteria (Figure 5F). Mice treated with AOH1996 had significantly longer survival (median 21 days) than vehicle-treated controls (or “excipients,” median 14 days, p=0.04). Mice in both groups had stable and similar weights during treatment (Figure 5G). Collectively, these studies provide evidence of preclinical efficacy and safety of TRC targeting using AOH1996 in PDAC.

### First evidence of AOH1996 efficacy in human metastatic PDAC

Given the promising preclinical activity, a phase 1 clinical trial was initiated with the primary goal of identifying maximum tolerated dose and dose limiting toxicity of AOH1996 in patients with refractory solid tumors. The first patient with PDAC enrolled on the trial derived clinical benefit from AOH1996. The patient presented with obstructive jaundice secondary to a mass in the head of the pancreas which was biopsy confirmed to be PDAC. The staging workup demonstrated metastases in the retroperitoneal lymph nodes and lungs. Genomic profiling demonstrated several mutations: KRAS(G12R), ARID1A (R1722*), and TP53 (S241F). The tumor was microsatellite stable, and tumor mutation burden was low. Germline testing was negative for pathogenic mutations. The patient was initiated on Gemcitabine and Nab-paclitaxel, to which there was stabilization of disease lasting 9 months. Subsequently, the patient was treated with 5-FU, leucovorin, and liposomal irinotecan and developed rapidly progressive disease within 3 months. The patient also developed two new liver metastases at this time. The patient was then enrolled on the phase 1 clinical trial (NCT05227326) and was treated with single agent AOH1996 (Dose level 3: 240 mg twice daily). After two months of therapy, the patient demonstrated stable disease in retroperitoneal lymph nodes and the primary tumor. Notably, there was approximately 49% tumor shrinkage in both hepatic metastases (Figure 6 A and B). However, there was slight growth of innumerable sub-cm lung metastases. Overall, the patient was deemed to have stable disease by RECIST (Response Evaluation Criteria in Solid Tumors) v 1.1. The patient continued AOH1996 for another 2 months, after which they demonstrated disease progression at all tumor sites and a rise in tumor marker CA 19-9 from 58 U/mL (pre-treatment) to 68 U/mL (post-treatment). Overall, this resulted in a progression free survival of 4 months on AOH1996. This case highlights proof-of-concept clinical activity of AOH1996 in this patient with chemotherapy refractory PDAC.

## DISCUSSION

Genome instability in PDAC is a function of endogenous and exogenous insults on the DNA. Endogenous mechanisms are now increasingly recognized to be a major driver of DNA damage and are a function of either deficient DNA repair pathways or oncogene-induced replication stress (29). While sporadic or germline DNA repair defects occur in up to 20% of PDAC patients; oncogene-induced replication stress is a uniform feature in PDAC – occurring in 95% of patients with KRAS driver mutations (30). The oncogenic program places significant demands on DNA replication mediated through anomalous cell growth signaling. Further, the oncogenic program also enhances the endogenous impediments to successful DNA replication that include: transcription machinery, torsional stress, and non-B DNA structures (27). Therefore, mechanisms that allow PDAC cells to replicate despite replication impediments may provide therapeutic opportunities.

Spatial and temporal deregulation of transcription and replication under aberrant oncogenic signaling can cause significant increase in TRCs (2,31,32). We have previously demonstrated that human PDAC demonstrates uniquely high levels (30-120-fold) of TRC-related mutational signatures compared to other common solid tumors (colon, breast and non-small cell lung cancer) (7). We have also established that TRCs in PDAC are a direct consequence of oncogenic KRAS activation and potentially represent a unique therapeutic vulnerability. The current study tests the overarching hypothesis that TRCs can be targeted for therapeutic purposes. The experiments presented here provide first evidence of therapeutic targeting of TRCs using a small molecule inhibitor in PDAC pre-clinical models as well as in a patient with metastatic PDAC.

The premise of targeting TRCs is based in the notion that cancer cells are particularly reliant on TRC avoidance and resolution mechanisms to sustain the extra-ordinary demand on replication and transcription. This reliance of cancer cells on TRC mechanisms therefore may create a therapeutic vulnerability that could allow for tumor-selective DNA damage. A growing body of literature provides pre-clinical evidence for TRC-targeting therapeutic approaches. For instance, BET bromodomain protein which are necessary for oncogenic transcription programs can be targeted using small molecule BET inhibitors (33). Bowry et. al demonstrated that BET inhibitors (specifically BRD4 inhibition) may enhance TRCs through enhanced transcription of highly transcribed histone and other non-poly-adenylated non-coding RNA genes. However, it remains to be defined if enhanced TRCs are related to the therapeutic effect of BET inhibitors (34). A recent report demonstrated that inhibition of Aurora A kinase in MYC-N amplified neuroblastoma can enhance transcriptional R-loops, TRCs, and ATR-checkpoint-driven DNA damage response. Combining Aurora A kinase and ATR inhibitor results in complete tumor regression in neuroblastoma mouse models (35). As another example, recently, MEPCE - a methylphosphate capping enzyme - was identified as a synthetic lethal interactor of BRCA1. Lack of MEPECE in a BRCA1-deficient context promoted RNAPII pausing, R-loops, TRCs, TRC-related DNA damage and reduced growth of breast cancer xenografts (36).

Distinct from the reported therapeutic approaches for targeting TRCs that enhance transcriptional pausing, AOH1996 demonstrates a unique trapping mechanism which promotes TRCs by enhancing the interaction of RNAPII and PCNA (19). Consistent with this trapping mechanism, we observed enhanced RNAPII-PCNA interaction in the PLA assay. Consequently, we note dose-dependent inhibition of replication fork progression due to AOH1996. Interestingly, inhibition of replication fork progression by AOH1996 was not associated with a significant compensatory increase in new origin firing, suggesting additional mechanisms of DNA replication inhibition that may impede replication licensing are at play. While AOH1996 may cause DNA damage by interfering with multiple other PCNA-related processes, our findings suggest that most of the AOH1996-related DNA damage in PDAC cells is likely a consequence of TRCs. The study uncovers a previously uncharacterized role of AOH1996 in causing transcription shutdown through proteosome-mediated degradation of transcriptionally active RNAPII. We speculate that AOH1996-mediated cancer cell death is realized through a combination of both TRC-induced DNA damage and transcription shutdown. Indeed, transcription inhibition through small molecule therapy (using triptolide) has previously been shown to be an effective strategy in PDAC models, albeit with high toxicity (37). In contrast, the lack of toxicity with AOH1996 in our studies may be related to tumor-selective transcription inhibition. Further ongoing studies will likely delineate additional mechanisms for therapeutic efficacy of AOH1996.

In PDAC organoid models we found considerable heterogeneity of response to AOH1996. Organoid models have been increasingly utilized to model clinical responses more accurately than conventional 2D models (38,39). For clinical translation, it is imperative to develop a biomarker of response to TRC-targeting approach. Dreyer et. al. first characterized a replication stress signature which was particularly enriched in the basal-subtype and did not overlap with DNA damage repair deficiency signature (18). Consistent with the observation of Dreyer et. al., we found overlap between basal and replication signatures. Further, we discovered that TRC-targeting was most effective in organoids exhibiting replication stress high or strongly basal transcriptomic signature. This is particularly relevant because basal-subtype is an aggressive variant of PDAC with the worst prognosis and limited response to the most effective chemotherapy (40). These observations provide a rationale for further development of replication stress high transcriptomic biomarker as a predictor of response to TRC-targeting therapies such as AOH1996 in the clinic.

Therapeutic responses in *in vitro* models were also recapitulated in the *in vivo* murine models. We noted a robust growth inhibition in both patient and mouse-derived models. In survival experiments, AOH1996 treatment prolonged survival but was not curative. While these observations provide a strong rationale for further development of a TRC-targeting strategy for PDAC, they also highlight the limitation in using monotherapy for successful clinical translation. Consistent with prior observations in mice and dogs (19), we found AOH1996 to be non-toxic in this study at effective doses. Notably, in this study, we provide first evidence of efficacy of a TRC-targeting approach in a metastatic PDAC patient who demonstrated shrinkage of hepatic tumors with AOH1996 monotherapy. However, we also noted heterogeneity of response with pulmonary metastases (demonstrating slight progression) and primary tumor (demonstrating stability). While longer in duration than prior systemic therapy, the response to AOH1996 was still short-lived with ultimate radiographic tumor progression. These early observations underscore the need not only to characterize optimal candidates for AOH1996 but also highlight the importance of studying the effect of AOH1996 in the context of tumor microenvironment. Ongoing work is exploring rational combination therapies in combination with TRC-targeting in PDAC.

## Supporting information

Supplemental Results

## Data Availability

All data produced in the present study are available upon reasonable request to the authors

## ACKNOWLEDGEMENTS

Research reported in this publication included work performed in the following Shared Resources supported by the National Cancer Institute of the National Institutes of Health under grant number P30CA033572: Analytical Cytometry; Light Microscopy & Digital Imaging; Integrative Genomics; Small Animal Imaging; and Research Histology. The content is solely the responsibility of the authors and does not necessarily represent the official views of the National Institutes of Health. This work was funded by the NCCN Foundation^®^. Any opinions, findings, and conclusion expressed in this material are those of the author(s) and do not necessarily reflect those of National Comprehensive Cancer Network^®^ (NCCN^®^) or the NCCN Foundation.

## METHODS

### Materials

AOH1996 was a generous gift from Dr. Long Gu and Dr. Linda Malkas and was synthesized and isolated to >95% purity in house by the Chemical GMP Synthesis Facility at City of Hope Comprehensive Cancer Center. Compound was received in powder form and stored at -20°C. Prior to use, for *in vitro* experiments, AOH1996 was dissolved in sterile dimethyl sulfoxide (DMSO, Fisher Scientific, Hampton, NH) to a concentration of 20 mM, and aliquots were stored at -20°C.

### Cell culture

All commercial cancer cell lines were cultured according to procedures established by the American Type Culture Collection (ATCC). UPN3 cell line was patient-derived and was a generous gift from Dr. Edward Manuel (City of Hope). MIA PaCa-2, PANC-1, KPC, and UPN3 cell lines were cultured in Dulbecco’s Modified Eagle Medium (DMEM) (Mediatech, Manassas, VA), supplemented with 10% fetal bovine serum (FBS) and 1% penicillin/streptomycin (P/S). Capan-1 cell lines were cultured in Iscove’s Modified Dulbecco’s Medium (IMDM) (Mediatech, Manassas, VA), supplemented with 20% FBS and 1% P/S. Capan-1 C2-14 cell line was a generous gift from Prof. Toshiyasu Taniguchi (41). BxPC-3 cell lines were cultured in RPMI-1640 (Mediatech, Manassas, VA), supplemented with 10% FBS and 1% P/S. All cell cultures were maintained at 37°C in 5% CO_2_. HPNE and iKRAS cells and their culture conditions have been described previously. iKRAS cell lines were a generous gift from Prof. Haoqiang Ying (25). All cells used in these studies were verified to be mycoplasma free within 6 months of the experiments. Organoid cell lines were generated from untreated patient resected tumors or biopsies as described previously (38).

### In vitro cytotoxicity of AOH 1996

Exponentially growing (1×10^3^ to 5×10^3^, depending on cell doubling time) pancreatic cancer cells (lines described above) were seeded in 96-well plates. In (at least) quadruplicate, increasing concentrations of AOH1996 were added to each well and incubated for 48 hours at 37°C in 5% CO_2_. Organoid experiments were performed in 384-well plates using a high-throughput protocol described previously (39). Organoids were exposed to control conditions or AOH1996 for 72 hours. Cell viability was determined using the CellTiter-Glo Luminescent Cell Viability Assay (Promega, Madison, WI), according to manufacturer’s instructions. Activity was calculated as % of cells alive/concentration versus control cells with no AOH1996 treatment, where 100% indicates no cell death (high ATP levels) and 0% indicates complete cell death (low or no ATP levels). Data were analyzed and IC_50_ values were determined following the guidelines described by Sebaugh et. al. (42) and using the sigmoidal dose-response equation in GraphPad Prism 5 software (La Jolla, CA).

### DNA Fiber Analysis

DNA fiber assays were performed using a modified version of the technique described by Frum et. al. (43). In brief, actively dividing MIA PaCa-2 cells (500,000 cells/well of a 6 well dish) were pulse-labeled with 100 µM of chlorodeoxyuridine (CldU) (Sigma-Aldrich, St. Louis, MO) for 15 minutes at 37°C and 5% CO_2_ in complete media. The CldU was subsequently removed from the cells by washing three (3) times with 1x phosphate buffered saline (PBS) (Corning, Tewksbury, MA). The cells were then pulse-labeled with 200 µM iododeoxyuridine (IdU) (Sigma-Aldrich, St. Louis, MO) for 30 minutes in the presence of 0, .5 µM, 1 µM, or 2 µM AOH1996 at 37°C, 5% CO_2_ for 30 minutes. After labeling, the cells were washed three (3) times with PBS and collected by trypsinization and centrifugation at 500xg for 5 mins. DNA spreads were prepared, and incorporated nucleosides were detected using IdU- and CIdU-specific antibodies. DNA fibers were imaged using a widefield light microscope at 100X magnification. (See Supplemental Methods for details).

### Western Blot Analysis

Western blot analysis was performed as previously described and noted in supplemental methods (21).

### Immunocytochemistry (ICC) Staining of γH2AX and Confocal imaging

Immunocytochemistry was performed as described previously (44) using standard protocol. For confocal imaging, a Zeiss LSM 700 Confocal Microscope (Jena, Germany) was used. Images were acquired using an LCI Plan Neofluar 63x/1.3 Water Imm Corr M27 objective for a 1024 x 1024- pixel array at 0.05 microns/pixel. (See Supplemental Information for details).

### Cell Proliferation Assay using Flow Cytometry

MIA PaCa-2 cells (either untreated or treated with 200 nM AOH 1996) were pulse-labeled with 10 µM of bromodeoxyuridine (BrdU) in DMEM supplemented with 10% FBS and 1% P/S for 1 hour at 37°C. Pulse-labeled cells were then recovered, washed, and processed for BrdU staining using the protocol specified in the BD Pharmingen BrdU Flow Kit (BD Life Sciences, San Jose, CA). Briefly, the cells were fixed, permeabilized, stained with anti-BrdU, and counter-stained with fluorescein isothiocyanate-conjugated (FITC) goat anti-mouse IgG1. Following 7-aminoactinomycin D (7-AAD) staining, cellular data was acquired using the BD FACSDiva software with the BD LSRFORTESSA (San Jose, CA). Offline analysis was performed using FlowJo v10 software (Ashland, OR, USA).

### Cell proliferation with Real-time cell analysis (RTCA)

For measuring the differential AOH1996 inhibitory effect on KRAS(G12D)-expressing cells, the RTCA assay was used to measure real-time cell growth. The HPNE-KRAS(G12D) cells were pretreated with or without 2 ng/ml Doxycycline for 72 hour until 60%-70% confluency; 4000 cells were then seeded on an E-plate (Agilent 05469830001). Cell growth curves were recorded on xCELLigence RTCA DP (Agilent, CA, USA) at 15-min intervals under standard cell culture conditions (37°C, 5% CO2). AOH1996 was added 40 hours after cells seeding at indicated concentrations.

### Cytofluorometric Analysis of Nuclear Apoptosis by TUNEL Assay

Nuclear apoptosis was assessed by the Terminal deoxynucleotidyl transferase (TdT) dUTP nick-end labeling (TUNEL) assay. This assay was performed using the In Situ Cell Death Detection Kit, TMR red kit (Roche, Basel, CH) according to the manufacturer protocol. Briefly, PBS-washed MIA PaCa-2 cells (2×10^7^ cells/ml) were fixed with 2% paraformaldehyde (MilliporeSigma, St. Louis, MO) for 60 minutes at 15-25°C. Fixed cells were washed twice in PBS and permeabilized using 0.1% Triton X-100 in 0.1% sodium citrate for 2 min on ice. After another PBS wash step, cells were incubated with 50 μl of TUNEL reaction mixture for 1 hr at 37°C in a dark, humidified atmosphere. TdT enzyme was not added for the negative control. Cells were washed twice more in PBS prior to analysis. Cellular data was acquired using the BD FACSDiva software with the BD LSRFORTESSA (San Jose, CA). Offline analysis was performed using FlowJo v10 software (Ashland, OR, USA).

### Proximity-ligation assay for TRC determination

To quantify TRCs we performed Proximity-ligation assay (PLA) as described previously (7). Briefly, treated cells at 60-70% confluence were trypsinized, washed at least one time with DPBS, then fixed with 4% PFA for 15 mins followed by permeabilization with 5-minute incubation with 0.5% NP-40. The fixed cell pellets were then collected after centrifugation at 400xg for 5 mins and washed with DPBS twice, blocked with commercial block buffer (supplied in kit) at room temperature for 1 hour, and then incubated with RNAPII-CTD S2 (1:100) and PCNA (1:100) antibody overnight in 4°C. PLA reactions were performed using the manufacturer supplied instructions in the kit (Sigma Aldrich). PLA-fluorescent labelled cells were passed through a 100µm strainer, and the PLA signal was measured using Attune NXT cytometer. FlowJo (Ashland, OR, USA) was used for data processing, analysis, and visualization.

### Global RNA transcription quantification

Global RNA quantification was performed using the Click-iT™ RNA Alexa Fluor™ 488 Imaging Kit (ThermoFisher Scientific, Waltham, MA, USA) according to manufacturer supplied methodology and as reported previously (45). Briefly, pancreatic cancer cells were plated at 40% confluence and allowed to attach overnight. The cells were then treated with indicated conditions for 48 hours. Subsequently, all cells were treated with 1mM 5-ethynyl uridine (EU) final concentration for 1 hour. Cell Fixation, Permeabilization, and Click-iT^®^ Detection were performed per protocol.

### RNA-seq Analysis

RNA sequencing was performed as described previously (39). The RNA-seq datasets are merged based on the Hg38 gene symbols. The extremely low expressed genes and the genes that were not detected in all datasets were filtered out. R-package ComBat-seq was used to remove the batch effects (46). Differential gene expression analysis was performed on the batch effect-corrected RNA-seq expression data using R-package DESeq2 ver 1.26.0 (47). The top differentially-expressed genes between drug resistant samples and drug sensitive samples were selected using adjusted p-value <0.05. The heatmap of differentially-expressed genes was generated using R-package pheatmap (ver 1.0.12). The principal component analysis (PCA) plot, the volcano plot and dot plot are generated using R-package ggplot2 (Ver 3.4). Basal and Classical classification and scoring was based on signatures that have been reported previously (28). Replication stress signature and scoring was generated based on a previous report (18).

### Mouse Models of Pancreatic Cancer

An orthotopic PDAC model was generated, as described previously (48,49). Approximately 1 million (1×10^6^) luciferase-expressing KPC cells were injected in the pancreas of a carboxyl esterase-deficient ES1^e^/SCID mouse using a 30-guage-needle tip (point style 4, Hamilton Company, Reno, NV). The KPC cell line was isolated from KPC (LSL-KrasG12D/+; LSL-Trp53R172H/+) mice that are known to recapitulate human pancreatic cancer. Tumor growth was monitored via bioluminescence imaging using a Lago scanner (Spectral Instruments Imaging Inc., Tucson, AZ, USA). Once tumors were confirmed (at approximately 2-3 weeks), mice were enrolled in the study, and treatments were assigned.

An ectopic patient-derived xenograft model was developed from a resected patient tumor that was passaged subcutaneously in immune-deficient NSG mice. Tumors from donor mice were harvested when they reached 1.5 cm in size. A 4 mm^2^ tumor fragment was placed in the recipient mice subcutaneously in the flank. Upon engraftment, mice were then randomized to AOH1996 or excipient groups.

After completion of treatment, mice were euthanized via carbon dioxide inhalation, and tumor weights were measured. All animals were handled, housed, and studied in accordance with a protocol (IACUC #18026) that was reviewed and approved by the City of Hope Institutional Animal Care and Use Committee. Office of Laboratory Animal Welfare, National Institutes of Health, U.S. Department of Health and Human Services guidelines were followed as well. (See Supplemental Methods for details).

### Immunohistochemistry on Mouse Tissues

Immunohistochemistry and TUNEL assay were performed using standard methods detailed in Supplemental Methods. For fluorescence immunohistochemistry, frozen tissues embedded in Optimal Cutting Temperature compound (OCT) were sectioned at 5 microns; and fixed on glass slides using acetone. Staining was performed similarly to immunocytochemistry. Confocal imaging was performed with a confocal microscope (Zeiss LSM 700, Jena, Germany) and analyzed using Image Pro Premier (version 9.3, Media Cybernetics, Inc., Rockville, MD). (Also see Supplemental Methods)

### Human studies

The patient treated with AOH1996 was enrolled under COH IRB# 21310 (NCT05227326) after informed consent. Contrast CT of the chest, abdomen, and pelvis performed from the lung apices through the pelvis was obtained before and after treatment at 2-month intervals per protocol. Tumor volume measurements were derived from contrast CT reconstructions as detailed in Supplementary Methods.

