## Supplemental Results for "Therapeutic Targeting of Oncogene-induced Transcription-Replication Conflicts in Pancreatic Ductal Adenocarcinoma"

**FIGURE S1**

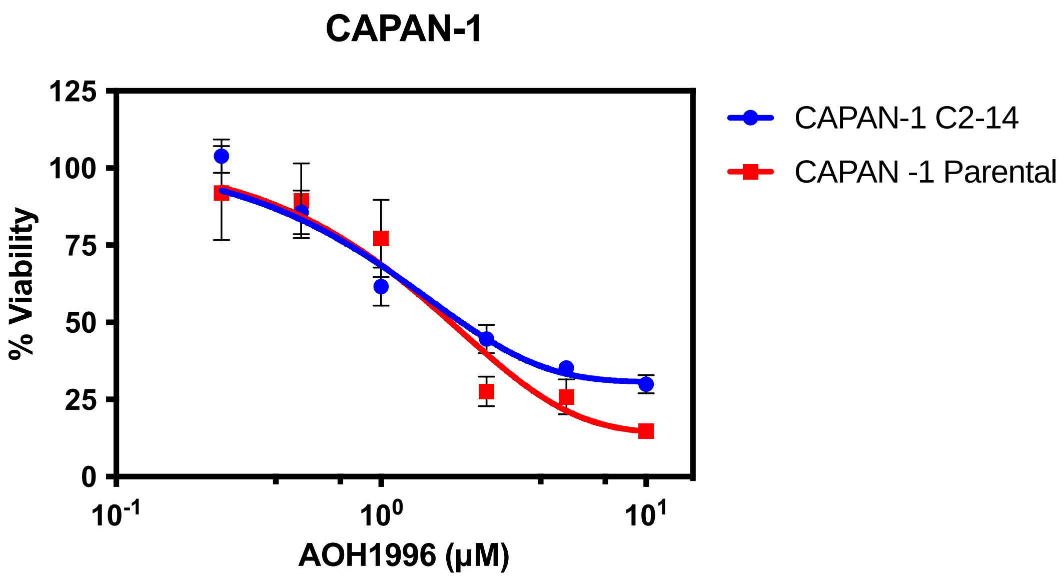


**Figure S1. Effect of AOH1996 on human CAPAN-1 viability using CellTiter Glo$^{TM}$ Assay**
Parental CAPAN-1 cells harbor pathogenic BRCA2 mutation (BRCA2.6174delT) that leads to homologous recombination repair deficiency, whereas C2-14 clone has a BRCA2 reversion that restores homologous recombination repair. AOH1996 demonstrated similar efficacy against the two cell lines. A representative experiment is shown with each datapoint representing mean of 4-6 observations and error bars indicating standard deviation. The line indicates non-linear best-fit values.

FIGURE S2

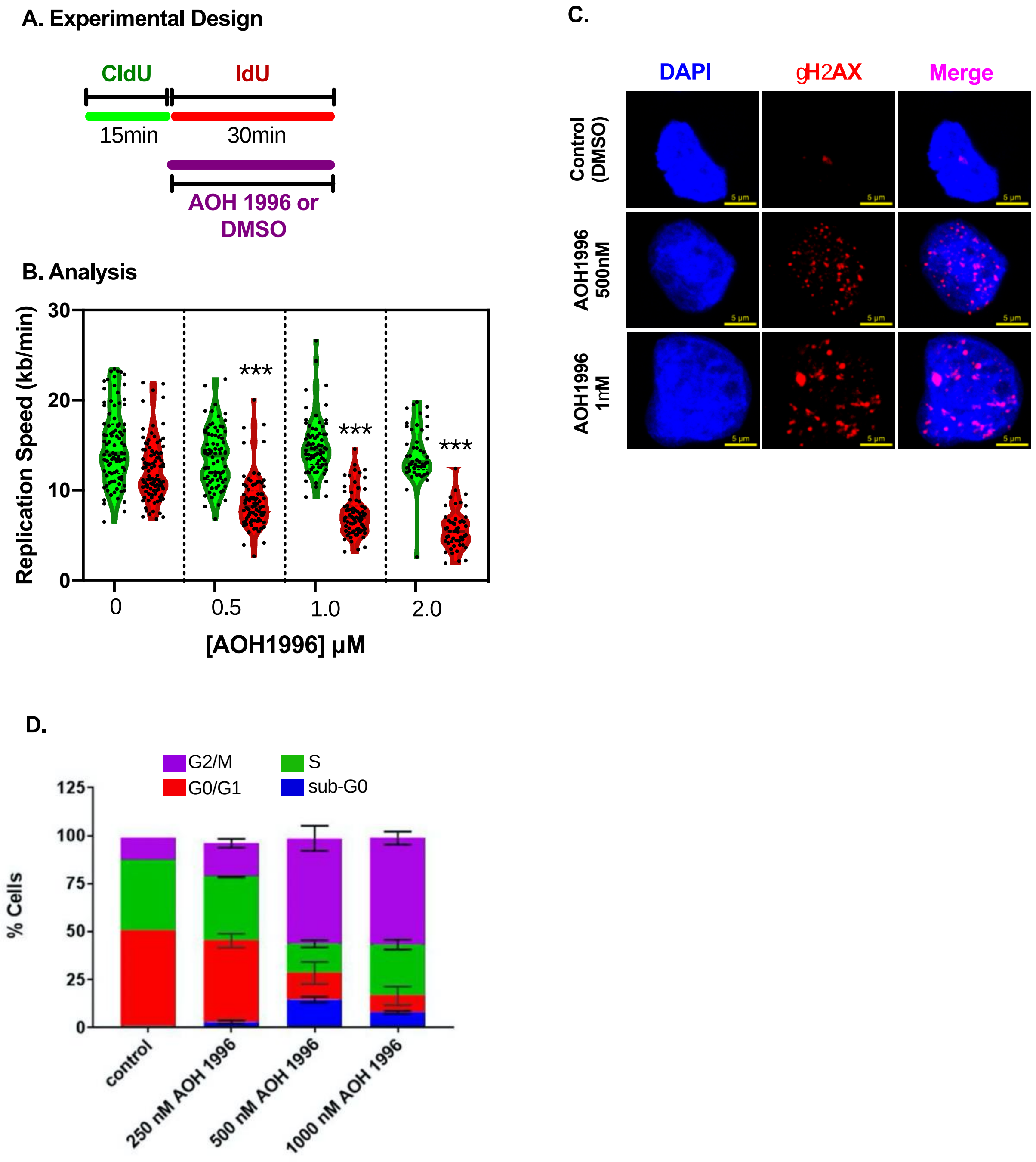


**Figure S2. Effect of AOH1996 on DNA replication in Pancreatic Cancer Cells.**

**A. Experimental Design.** DNA fiber analysis was performed. Replicating DNA was labeled with CldU for 15min, followed by IdU for 30min. AOH 1996 or DMSO was added with IdU labeling.

**B. Analysis:** CldU and IdU tract lengths were measured (in microns) at each concentration of AOH1996 and were normalized to label exposure time to calculate replication speed. (1 micron = 2.59 kilobases). One-way ANOVA demonstrated significant decrease in replication speed during AOH1996 exposure (***p<0.0001 for Dunnett multiple comparison test. Comparison performed with Red Violin Plot at 0μM). **C. Immunocytochemistry:** Representative images for DNA damage marker (gH2AX) for the experiment shown in Figure 2E. **D. Flow cytometry** analysis similar to experiment shown in Figure 2F & 2G. The dose dependence of cycle changes were recorded after 24 hours of AOH1996 exposure, and then analyzed in triplicates. Average proportions are plotted in stacked bar graph with error bars indicating standard deviation.

**FIGURE S3**

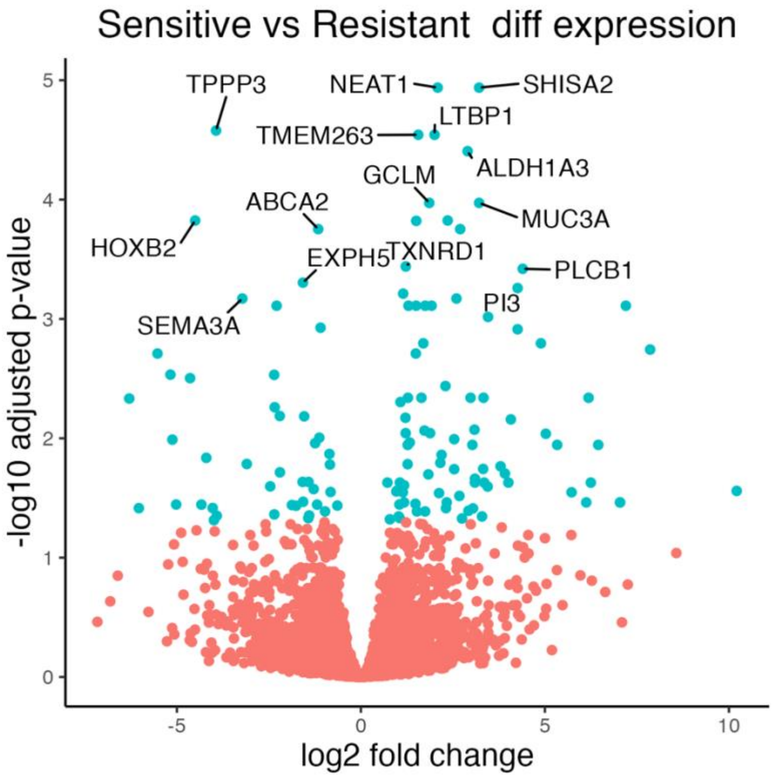


**Figure S3 Volcano plot demonstrating differential gene-expression between resistant and sensitive organoid lines.** The plot highlights the genes that are significantly differentially expressed in blue (Adjusted p-value <0.05). The top 20 differentially expressed genes based on the adjusted p-value are labeled.

**FIGURE S4**

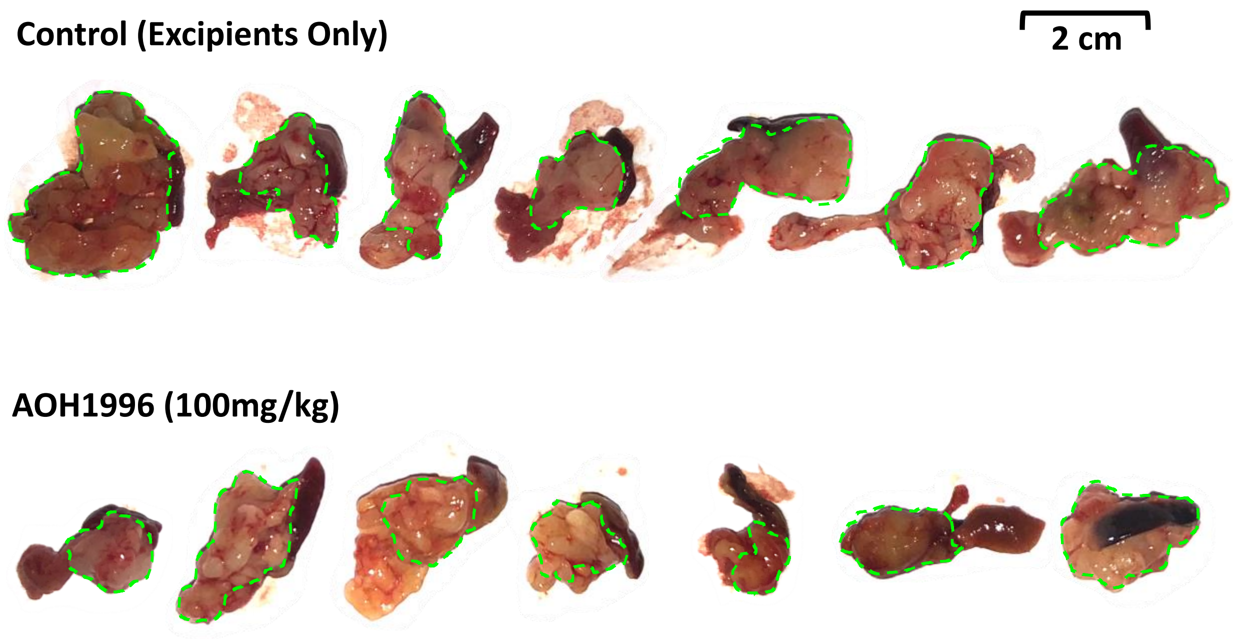


**Figure S4. Photograph of Pancreatic Tumors Harvested from Experimental Mice (Figure 5A).** Green outline identifies the area of the tumor

**FIGURE S5**

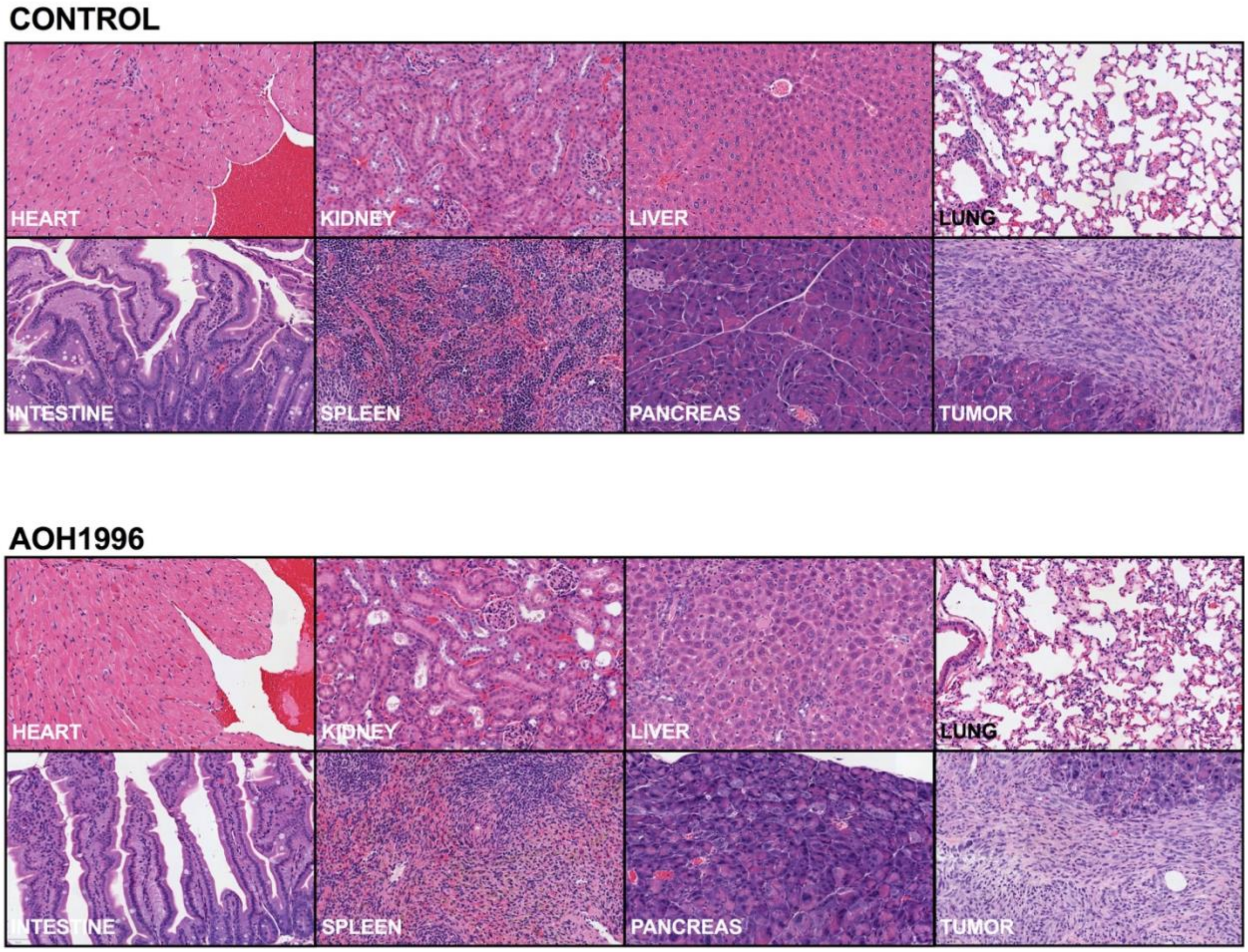


**Figure S5. Representative H&E images from tumor and normal tissues from the control group vs. AOH1996 treated mice.** No toxicity was observed on histologic analysis.

**FIGURE S6**

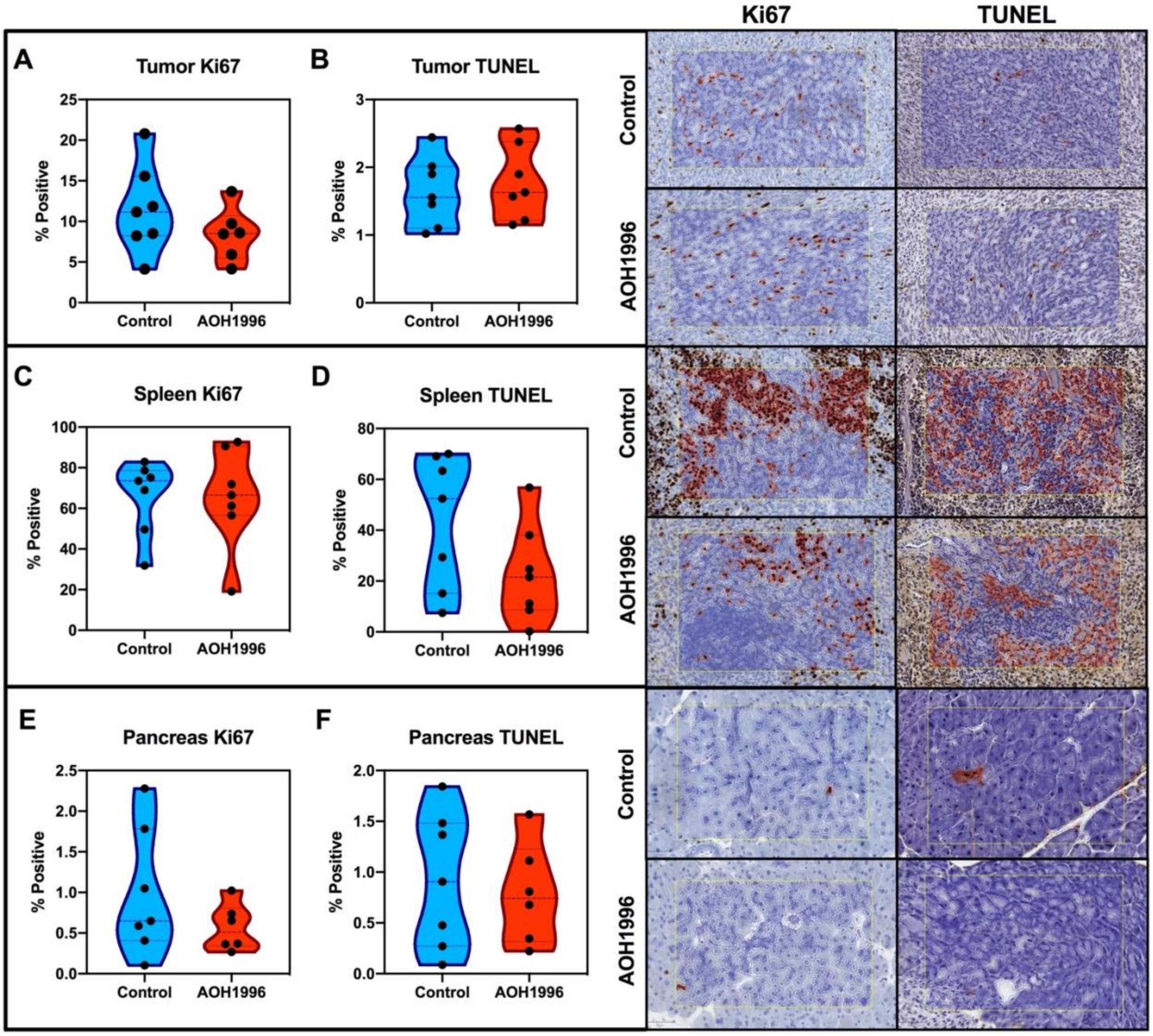


**Figure S6. Immunohistochemistry in mouse tumors and normal tissues**

There were minimal changes in proliferation index (determined by Ki67) and apoptosis (determined by TUNEL staining) of mouse tumor and normal tissues. These changes were not statistically significant (t-test, $p>0.05$ for all comparisons).

**SUPPLEMENTAL METHODS**

### DNA Fiber Analysis

DNA fiber assays were performed using a modified version of the technique described by Frum et. al. (1).In brief, actively dividing MIA PaCa-2 cells (500,000 cells/well of a 6 well dish) were pulse-labeled with 100 µM of chlorodeoxyuridine (CldU) (Sigma-Aldrich, St. Louis, MO) for 15 minutes at 37$^oC$ and 5% $CO_2$ in complete media. The CldU was subsequently removed from the cells by washing three (3) times with 1x phosphate buffered saline (PBS) (Corning, Tewksbury, MA). The cells were then pulse-labeled with 200µM iododeoxyuridine (IdU) (Sigma-Aldrich, St. Louis, MO) for 30 minutes in the presence of 0, .5 µM, 1 µM, or 2 µM AOH1996 at 37$^oC$, 5% $CO_2$ for 30 minutes. After labeling, the cells were washed three (3) times with PBS and collected by trypsinization and centrifugation at 500xg for 5 mins. The pelleted cells were resuspended in PBS and counted with a Beckman Coulter Z2 particle counter (Brea, CA). Two thousand cells were spread on a microscope slide (Leica, Wetzlar, Germany) and lysed by layering lysis buffer (0.5% SDS, 200 mM Tris-HCL pH 7.4, 400mM NaCl, 0.2% NP40) on the cells. The slides were placed at an angle (15°-45°) to allow the DNA fibers to spread down the slides. Four replicate slides were made for each experimental condition. The slides were then fixed with 3:1 methanol/acetic acid and the DNA denatured with 2.5M HCl. Following a blocking step, the DNA fibers were hybridized with an antibody specific to CldU (Abcam, Cambridge, UK) that was derived from rat, and an antibody specific to IdU (BD Biosciences, San Jose, CA) that was derived from mouse. The primary antibodies were then detected using secondary antibodies conjugated to a fluorophore: goat anti-rat IgG conjugated to AlexaFluor 488 (Thermo Fisher Scientific, Waltham, MA) and rabbit anti-mouse IgG conjugated to AlexaFluor 594 (Thermo Fisher Scientific, Waltham, MA). After washing, coverslips were mounted onto slides and DNA fibers were imaged using the fluorescent setting of the Zeiss Observer II (Carl Zeiss AG, Oberkochen, DE) widefield light microscope at 100X magnification. In post-processing, Image J (NIH, Bethesda, MD) was used to visualize and measure green and red fluorescing lengths of DNA fibers. The fibers chosen for measurement were those with a clearly defined section of a green fluorescence followed immediately by a section of red fluorescence, and only fibers with similar lengths of green fluorescence (as judged typical for the experiment) were scored.

### Western Blot Analysis

Western blot analysis was performed as previously described (2). Cells were seeded in 100 mm tissue culture treated culture dishes, grown to approximately 75% confluency, and treated with 200 nM AOH 1996. Treatment was stopped at specified time points (4, 24, and 48 hours), and cells were washed three times in ice cold TBS (20 mM Tris, pH 7.6, and 137 mM NaCl). The cells were then harvested into TBS, 2x ThermoScientific Halt phosphatase inhibitor cocktail, 2x Halt protease inhibitor cocktail (Thermo Fisher Scientific, Waltham, MA), and 10 mM EDTA. Harvested cells were pelleted in a swinging bucket centrifuge at 1,500 rpm for 5 min, the supernatant was removed, and the pellets were stored at -80 $^oC$ prior to processing. Thawed pellets were then sonicated in SDS buffer (100 mM Tris, pH 6.8, 4% SDS, 20% glycerol, 1x ThermoScientific Halt phosphatase inhibitor cocktail, 1x Halt protease inhibitor cocktail, and 5 mM EDTA) at 30%

amplitude in 10 sec intervals until no longer viscous. After heating at 95 $^{o}$C for 5 min and cooled, the total protein concentration was determined by DC Protein Assay (Bio-rad, Hercules, CA). DTT and bromophenol blue was added to a final concentration of 90 mM DTT and ~0.08% bromophenol blue, respectively. After resolving the protein extract by SDS PAGE, it was transferred to nitrocellulose membrane using the Pierce G2 Fast Blotter (Thermo Fisher Scientific, Waltham, MA) and total protein loading was visualized using Ponceau-S staining solution (Sigma-Aldrich, St. Louis, MO). The blots were blocked in 5% nonfat dried milk and 0.05% tween 20 for 1 h. Antibodies recognizing H2AX (Cell Signaling Technology, Beverly, MA, USA), phospho-histone H2AX Ser139 (Millipore, Burlington, MA, USA), RNAPII (A10, Santa Cruz Biotech, Santa Cruz, CA, USA), and actin (Abcam, Cambridge, UK) were detected by ECL prime and imaged on the Azure c600 (Azure Biosystems, Dublin, CA, USA).

**Immunocytochemistry (ICC) Staining of γH2AX**

Immunocytochemistry was performed as described previously (3). Circular #1.5 cover slips (Electron microscopy sciences, Hatfield, PA) were placed in 12-well plates and sterilized by UV exposure for 20 minutes. Approximately 15,000 exponentially growing MIA PaCa-2 cells were seeded in each well, and adherent sub-confluent monolayers were observed growing on the cover slip 24 hours later. Cells were then exposed to various treatment conditions, as described in the Results section. After treatment, cells were fixed in 1% paraformaldehyde in PBS (w/v) for 30 minutes, permeabilized using 0.3 % (v/v) Triton-100 and 0.125 % (w/v) CHAPS (3-[(3-Cholamidopropyl) dimethylammonio]-1-propanesulfonate) dissolved in PBS for 15 minutes, blocked for 1 hour in a 3 % (w/v) bovine serum albumin and 1 % (v/v) normal goat serum, labeled with γ-H2AX mouse monoclonal primary antibody (Millipore, Burlington, MA), followed by secondary labelling with Alexa Fluor 647-conjugated goat anti-mouse antibody. At the end of immunolabeling, 4',6-diamidino-2-phenylindole (DAPI) (Millipore, Burlington, MA) was used to counter stain DNA at 0.1 µg/mL for 15 minutes. Between each step, cells were washed with PBS three times for 5 minutes each time on a leveled shaker at 50 RPM. Cover slips were washed one more time with PBS and were mounted on frosted glass slides (Thermo Fisher Scientific, Waltham, MA) using Dako mounting media (Dako, Carpinteria, CA). The slides were sealed with a conventional nail polish hardener and stored at 4$^{o}$C until imaging.

**Confocal Imaging**

For confocal imaging, a Zeiss LSM 700 Confocal Microscope (Jena, Germany) was used. Images were acquired using an LCI Plan Neofluar 63x/1.3 Water Imm Corr M27 objective for a 1024 x 1024-pixel array at 0.05 microns/pixel. Samples were excited using a solid-state laser at 405nm (768 gain) for DAPI, 639nm (901 gain for the first biological replicate and 777 gain for the second biological replicate) for Alexa Fluor 647. Exposure settings were set to maximize dynamic range initially and then kept constant across multiple samples to allow quantitative comparisons. Acquired images were processed in Image Pro Premier (version 9.2, Media Cybernetics, Inc., Rockville, MD). Nuclei were identified using the DAPI channel. For counting γ-H2AX foci, as there was background signal for the Alexa Fluor 647 signal, the images from the control condition (DMSO treatment) were used to determine a threshold for the Alexa Fluor 647 signal, defining the threshold such that less than 10% of cells in the control condition were counted as positive for γ-H2AX foci, and signals that appeared to be adjacent foci were counted as individual foci rather than counting two foci as one. Positive cells were defined as those having five or more γ-H2AX foci in the nucleus. For each biological replicate, the threshold was determined using the control condition within that replicate, and this threshold was applied to all images acquired as part of that replicate; for the first biological replicate, the Alexa Fluor 647 signal intensity threshold was set to the range 58-255 (arbitrary units); for the second biological replicate, the threshold was set to the range 98-255 (arbitrary units). For foci segmentation purposes, Smoothing was set to 1, and Grow was set to -1, and Watershed splitting was applied during foci counting. Approximately 100 cells per condition per experiment were counted.

**Immunohistochemistry (IHC)**

IHC was performed on Ventana Discovery Ultra IHC autostainer (Ventana Medical Systems, Roche Diagnostics, Indinapolis, USA) using the ChromoMap DAB detection system according to manufacturer's recommendations. Briefly, tissue samples were sectioned at a thickness of 5 µm and put on positively charged glass slides. Deparaffinization, rehydration, endogenous peroxydase activity inhibition and antigen retrieval were all performed on the automated stainer. Slides were then incubated with either primary rabbit anti-K67 monoclonal antibody (Ventana), or rabbit anti-H2AX monoclonal antibody (abcam), followed by DISCOVERY anti-rabbit HQ and DISCOVERY anti- HQ-HRP, visualized with ChromoMap DAB detection Kit (Ventana). The slides were then counterstained with haematoxylin (Ventana) and coverslipped.

**TUNEL on Mouse Tissues**

To determine apoptosis, deoxynucleotidyl transferase (TdT)-mediated dUTP-biotin nick end labelling (TUNEL) method was performed by using The ApopTag Peroxidase In Situ Apoptosis Detection Kit (Millipore, Burlington, MA) based on the manufacturer’s instructions. Briefly, Sections were deparaffinized and washed in PBS, followed by proteinase K pretreatment and Endogenous Peroxidase activity quenching. Labeling was performed by adding TdT enzyme mix to the tissue sections and the reaction was stopped by immersing slides in stop buffer followed by three PBS washes. After adding anti-digoxignenin conjugate to the slide; the color was developed by Peroxidase Substrate Kit diaminobenzidine (DAB) (Vector Laboratories, Inc.). Finally, the slides were counterstained with haematoxylin (Ventana) and coverslipped.

**Image Acquisition and Quantification for IHC**

All glass slides were digitized in a VENTANA iScan HT (Roche-Ventana Medical Systems, Tucson, AZ, USA) at a magnification of ×40. Quantification profiles of the digitized whole slides were performed using QuPath software (4).Quantification of the positive cells with the Ki67 and TUNEL stains was carried out with the positive cell detection function. To avoid selection bias and increase statistical accuracy, the entire tissue section was manually selected using the polygon tool taking care to exclude areas rich in immune and stromal cells. The software performed annotation and quantification and expressed the positive cell counts as a percentage of positive cells detected per total cells.

**Tumor Implantation**

Male and female mice between 17-18 weeks in age were used in the study. Before surgery, hair was removed from the left side of the torso using clippers. Mice were anesthetized with 5% isoflurane initially and were kept anesthetized with 2% isoflurane during surgery. The surgical field was sterilized with Clidox. Mice were placed lying on their right side on a heating pad. The surgical site was sterilized with alcohol and povidone iodine, and sterility was maintained, including using a drape for protection. An abdominal incision left of the midline was made to expose the pancreas. A volume of 10 µL of KPC cells was injected into the pancreas using a Hamilton syringe with a 30-guage-needle tip (point style 4, Hamilton Company, Reno, NV). The pancreas was re-internalized, and incised tissue was closed using a suture and using wound clips (Fine Science Tools, Foster City, CA). Mice were allowed to regain consciousness in a cage placed halfway on a heating pad and under a thermal lamp and were observed for 10-20 minutes before returning them to housing. Mice were given buprenorphine SR as an analgesic after allowing them to awaken from anesthesia. Wound clips were removed 12-14 days after surgery.

**Bioluminescence Imaging**

Two to three weeks after implantation of tumor cells in the pancreas, bioluminescence measurements were performed to confirm the presence of tumor in the pancreas. D-Luciferin from firefly (Gold Biotechnology) was administered intra-peritoneally (i.p.) at a dose of 200 ug in 200 uL PBS. Animals were anesthetized using 2% isoflurane and imaged using a Lago scanner (Spectral Instruments Imaging Inc., Tucson, AZ, USA) 10-12 minutes after the injection. The imaging was performed with an exposure time of 10 seconds with 2-fold binning, a 1.2 f/stop, and a 25 cm field of view. Mice that had any bioluminescence activity above background (suggesting the development of tumors) were included in the study.

**Treatment Assignment and Administration**

Mice with confirmed tumors based on bioluminescence were then randomized into two groups, either treatment with AOH1996 or treatment with vehicle control using the Random.org List Randomizer (https://www.random.org/lists/). Randomization was blinded as the tumors were not visible to naked eye and the investigator performing the randomization was separate from the investigator administering treatments.

Mice with tumors were treated once per day for four days with either 100 mg/kg AOH1996 or vehicle control via oral gavage. The formulation was previously described (Gu, et al., 2018); this formulation allowed for a concentration of AOH1996 at 22.6 mg/g. The formulation was mixed at 60% with sterilized ultrapure water (40%) and sonicated until homogenous; this step was performed immediately before treatment. After four days of treatment, mice were euthanized using dual method. Necropsy was performed and tumor masses were measured. Tumors, normal pancreas, spleen, lung, heart, intestine, kidney and livers were harvested for downstream studies.

**Image acquisition**

Contrast CT of the chest, abdomen, and pelvis performed from the lung apices through the pelvis. Images of the abdomen obtained before and after the administration 125 mL Isovue-370 contrast. Postcontrast images of the chest and pelvis obtained. Oral contrast was administered. Coronal and sagittal reformatted images obtained. Up-to-date CT equipment and radiation dose reduction techniques were employed. CTDIvol: 5.9 - 6.1 mGy. DLP: 341 mGy-cm.

Initial images were reconstructed into 0.625 x 1.25 mm slices for 3D post-processing and uploaded from the picture archiving and communication system (PACS) to advanced imaging software (Vitrea Advanced Visualization 7.15.2) for volumetric analysis.

**3D Volumetric measurement:**

Contrast CT images of the abdomen were transferred into FDA approved Vital Images advanced imaging workstation (Vitrea Advanced Visualization 7.15.2). All thin reconstructed images were uploaded into the 3D workstation. The Liver and both tumor volumes were extracted with the automated mode or by drawing the region of interest (ROI) around the tumor. The portal vein, Hepatic vein, and Hepatic Artery segmentation were performed using the automated tool. All 2D Images were then edited with the manual tool (as required on each slice) on specific slices and analyzed for liver and liver tumor volume quantification. Changes in the volume will be assessed by using the density-based voxel quantification which includes liver volume, tumor volume, average, minimum, and maximum in the Hounsfield unit. All the numbers are saved in Excel file for the purpose of statistical analysis.
